# Cognitive and inflammatory heterogeneity in severe mental illness: Translating findings from blood to brain

**DOI:** 10.1101/2023.11.02.23297924

**Authors:** Linn Sofie Sæther, Attila Szabo, Ibrahim A. Akkouh, Beathe Haatveit, Christine Mohn, Anja Vaskinn, Pål Aukrust, Monica B. E.G. Ormerod, Nils Eiel Steen, Ingrid Melle, Srdjan Djurovic, Ole A. Andreassen, Torill Ueland, Thor Ueland

## Abstract

Recent findings link cognitive impairment and inflammatory-immune dysregulation in schizophrenia (SZ) and bipolar (BD) spectrum disorders. However, heterogeneity and translation between the periphery and central (blood-to-brain) mechanisms remains a challenge. Starting with a large SZ, BD and healthy control cohort (*n*=1235), we aimed to i) identify candidate peripheral markers (*n*=25) associated with cognitive domains (*n*=9) and elucidate heterogenous immune-cognitive patterns, ii) evaluate the regulation of candidate markers using human induced pluripotent stem cell (iPSC)-derived astrocytes and neural progenitor cells (*n*=10), and iii) evaluate candidate marker messenger RNA expression in leukocytes using microarray in available data from a subsample of the main cohort (*n*=776), and in available RNA-sequencing deconvolution analysis of postmortem brain samples (*n*=474) from the CommonMind Consortium (CMC). We identified transdiagnostic subgroups based on covariance between cognitive domains (measures of speed and verbal learning) and peripheral markers reflecting inflammatory response (CRP, sTNFR1, YKL-40), innate immune activation (MIF) and extracellular matrix remodelling (YKL-40, CatS). Of the candidate markers there was considerable variance in secretion of YKL-40 in iPSC-derived astrocytes and neural progenitor cells in SZ compared to HC. Further, we provide evidence of dysregulated RNA expression of genes encoding YKL-40 and related signalling pathways in a high inflammatory subgroup consisting predominantly of SZ in the postmortem brain samples. Our findings suggest a relationship between peripheral inflammatory-immune activity and cognitive impairment, and highlight YKL-40 as a potential marker of cognitive functioning in a subgroup of individuals with severe mental illness.

## Introduction

Cognitive impairment is a major contributor to functional disability in severe mental illnesses (SMI), including schizophrenia (SZ) and bipolar (BD) spectrum disorders (Green, 2006; Wingo et al., 2009). Existing pharmacological treatments have little to no effect on cognitive symptoms (Nielsen et al., 2015; Senner et al., 2023), which may precede illness onset (Dickson et al., 2012; Sheffield et al., 2018) and persist throughout the illness course (Bonner-Jackson et al., 2010; Flaaten et al., 2023, 2022). While several pathophysiological mechanisms may be involved (McCutcheon et al., 2023), inflammatory and immunological dysregulation observed in SMI have emerged as a possible contributor to cognitive impairment (Kogan et al., 2020; Misiak et al., 2018; Morozova et al., 2022; Rosenblat et al., 2015). Inflammatory and immune-related signals may impact cognitive functioning through communication between the immune- and central nervous systems (Dantzer, 2018; Kronfol and Remick, 2000; Pape et al., 2019; Salvador et al., 2021). Increasing our mechanistic understanding of inflammatory-immune involvement may enable the development of novel treatment strategies for cognitive impairment across brain disorders including SMI.

Genetic, postmortem and epidemiological studies support the role of immune-mediated activity in the pathophysiology of SMI (Andreassen et al., 2023; Benros et al., 2014; Webster, 2023). Dysregulated levels of peripheral inflammatory and immune-related markers and mediators are apparent at all stages of illness (Goldsmith et al., 2016; Halstead et al., 2023; Perry et al., 2021; Upthegrove et al., 2014). Comorbid cardiometabolic disease may contribute to enhanced peripheral inflammation and immune activation in SMI (Foiselle et al., 2022; Zhang et al., 2022), although several markers remain altered independent of comorbidity (Mørch et al., 2019). Dysregulated peripheral levels may also arise from and contribute to neuroinflammation induced by immunocompetent glial cells (Almeida et al., 2019; Bishop et al., 2022), including disruption of the blood-brain barrier (BBB, Futtrup et al., 2020; Lizano et al., 2023b), or the blood-cerebral spinal fluid barrier (Bitanihirwe et al., 2022).

Case-control studies may mask clinically relevant characteristics of subgroups of individuals with SMI. Recent studies using a comprehensive selection of inflammatory and immune-related markers in SMI have revealed considerable variation in the degree of dysregulation (Bishop et al., 2022; Boerrigter et al., 2017; Enrico et al., 2023; Fillman et al., 2016, 2014; Hoang et al., 2022; Lizano et al., 2023a, 2020; Sæther et al., 2023; Tamminga et al., 2021; Zhang et al., 2022). A consistent finding is that 30-50% of individuals with SMI show inflammatory-immune dysregulation (Bishop et al., 2022; Boerrigter et al., 2017; Lizano et al., 2023a, 2020; Zhang et al., 2022), which is associated with several unfavorable outcomes, including cognitive impairment (Fillman et al., 2016; Lizano et al., 2023a, 2020). Similarly, there is evidence of transdiagnostic cognitive subgroups across SZ and BD spectrum disorders related to several negative outcomes (Carruthers et al., 2019; Vaskinn et al., 2020; Wenzel et al., 2023). Cognitive subgroups have also been associated with higher inflammatory and immune-related activity (Pan et al., 2020). These findings may underlie the weak associations that have been reported between inflammatory-immune markers and cognitive domains in SMI (Morrens et al., 2022). However, we still lack a clear understanding of the interplay between peripheral and central inflammatory-immune activity, and its impact on cognitive functioning in SMI (Khandaker and Dantzer, 2016; Tomasik et al., 2023).

Individuals with SMI and high inflammatory-immune dysregulation have abnormalities in brain structure and function, overlapping with cognitive impairment (Goldsmith et al., 2023; Hoang et al., 2022; Lalousis et al., 2023; Lizano et al., 2023a, 2020). These alterations may result from pathophysiological changes, as suggested by evidence using human induced pluripotent stem cell (iPSC) technology (Akkouh et al., 2021, 2020; Falk et al., 2016; Lizano et al., 2023b; Sheikh et al., 2022; Szabo et al., 2021). Using this method, skin cells are reprogrammed into iPSCs, which can be differentiated into a variety of central nervous system (CNS) cells. We have previously shown glia-related functional changes and abnormal secretion of inflammatory-immune markers in iPSC-derived astrocytes from SMI donors, both at resting state and in response to inflammatory challenge (Akkouh et al., 2021, 2020; Sheikh et al., 2022; Szabo et al., 2021). Inflammatory and immune-related markers and mediators secreted by the CNS may have neuromodulatory effects (Salvador et al., 2021), and emerging evidence highlights astrocyte signaling in neural processes supporting cognition (Santello et al., 2019). Identifying peripheral markers sharing variance with cognitive phenotypes, rather than diagnostic categories, in combination with investigating the regulation of these molecules in CNS cells using iPSC-technology, may improve our understanding of their pathogenic role in cognitive functioning in SMI. We recently identified transdiagnostic subgroups based on the shared variance between peripheral inflammatory-immune pathways and cognitive functioning, particularly measures of verbal learning and processing speed (Sæther et al., 2023). The current study aimed to use a translational approach to shed light on the relationship between peripheral and central inflammatory-immune activity related to cognitive functioning in SMI.

We first investigated patterns of covariance between cognitive functioning and a range of inflammatory-immune markers and mediators, followed by hierarchical clustering to capture heterogeneity across diagnostic categories and healthy controls in our main sample (SZ=435, BD=218, healthy controls, HC=582). We included nine core cognitive domains: fine-motor speed, psychomotor processing speed, mental processing speed, attention, verbal learning, verbal memory, semantic fluency, working memory and cognitive control. While we previously investigated markers reflecting BBB integrity and trafficking, chemokines, cell adhesion molecules and mediators of innate immune activity (Sæther et al., 2023), we now included a novel set of markers capturing a more comprehensive range of peripheral inflammation and immune activity reflecting i) immune activation of different leukocyte subsets, ii) general upstream inflammatory markers, iii) vascular and endothelial-related inflammation, iv) extracellular matrix (ECM) remodelling, v) Wnt signalling pathway and vi) neuroinflammation. Following identification of candidate markers sharing variance with cognitive domains, we assessed the potential dysregulation of these candidates in CNS cells, by *in vitro* inflammatory modulation of iPSC-derived astrocytes and neural progenitor cells (NPCs). The potential contribution of immune cells to systemic protein levels was evaluated by microarray analysis of mRNA levels in a sub-sample of the main cohort (n=776). Finally, dysregulation of candidate markers in the brain in relation to diagnosis and inflammatory sub-groups was assessed by RNA-sequencing and deconvolution analysis of postmortem brain samples (n=474) from the CommonMind Consortium (CMC). See Fig. 1 for overview of samples and analysis plan.

**Figure 1.**
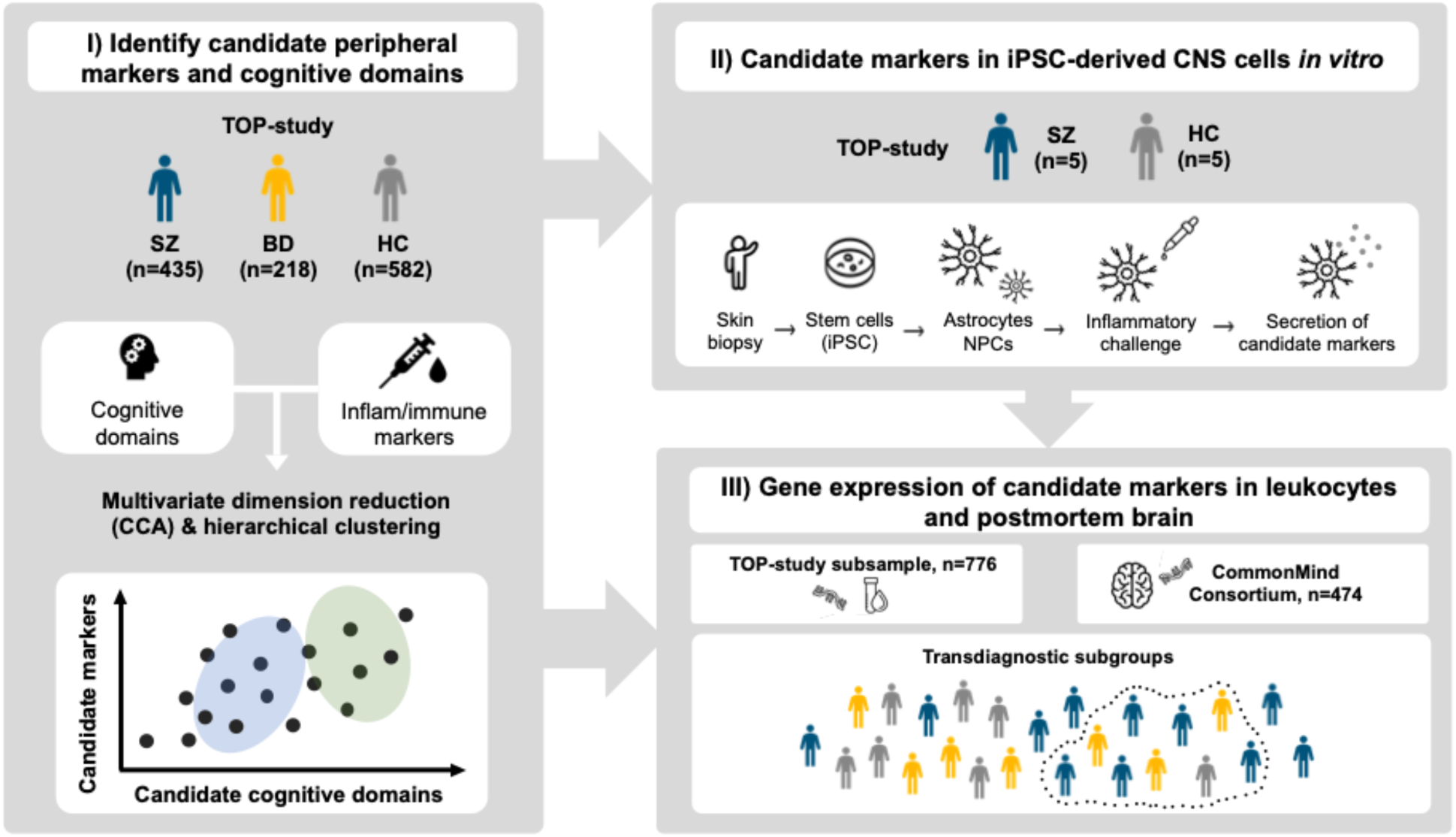
Study overview of samples and analysis plan.

## Methods

### Samples

This study is part of the ongoing Thematically Organized Psychosis (TOP)-study, with data from the main sample collected between 2003-2013. The main sample consisted of participants meeting the Diagnostic Manual of Mental Disorders (DSM)-Ⅳ criteria for severe mental illness, including schizophrenia (*n*=435), and bipolar (*n*=218) spectrum disorders were included in this study, in addition to healthy controls (*n*=582). Exclusion criteria were: 1) age <18 or >65, 2) moderate/severe head injury, 3) severe somatic/neurological disorder, 4) not fluent in a Scandinavian language, 5) IQ<70, 6) sign of acute infection (C-Reactive Protein>20 mg/L). Healthy controls were excluded in the case of drug abuse, history of mental illness, or relatives with SMI. Skin biopsies (fibroblasts) available from SZ (*n*=5) and HC (*n*=5) in the TOP-study were selected based on confirmed diagnosis/HC category and availability of a comprehensive set of data was used for astrocyte and NPC differentiation (see Supp. Table 1). For evaluation of leukocyte mRNA expression, a subcohort of the main sample with available data was included (SZ=311, BD=156, HC=309). All participants provided informed consent and the study was approved by the Regional Ethics Committee. In addition, we included an external sample of postmortem brain data (SZ=214, BD=45, HC=215) obtained from the CMC (Hoffman et al., 2019).

### Clinical assessments

Trained psychologists or physicians administered the Structured Clinical Interview for DSM-Ⅳ axis 1 disorders (SCID-Ⅰ) (First et al., 1995). Positive and negative symptoms were assessed using the Positive and Negative Syndrome Scale (PANSS) reported according to the five-factor model (Kay et al., 1987; Wallwork et al., 2012), and manic symptoms with the Young Mania Rating Scale (YMRS) (Young et al., 1978). The split version of the Global Assessment of Functioning scale (GAF-S, GAF-F) assessed functioning (Pedersen et al., 2007). Age at onset (AAO) was defined as the age of the first SCID-verified psychotic episode (SZ) or manic/hypomanic episode (BD). Duration of illness (DOI) was defined by subtracting the AAO from age at inclusion. Clinical interviews, physical examination (blood sampling, height/weight), and cognitive assessment occurred within 60 days. The defined daily dose (DDD) of psychopharmacological treatment (antipsychotics, antidepressants, antiepileptics and lithium) was determined according to World Health Organization guidelines (https://www.whocc.no/atc_ddd_index). Somatic medication use (including anti-inflammatory/immunomodulatory; yes/no) in the SMI group is provided in Supp. Table 2.

### Cognitive assessments

Cognitive assessment was undertaken by trained psychologists or research personnel using one of two test batteries: Battery 1 (from 2003-2012) or Battery 2 (from 2012). To ensure the highest possible N, corresponding tests from the two batteries were standardized separately (Z-scores) before collapsing to cover nine cognitive domains: *Fine-motor speed, psychomotor processing speed, mental processing speed, attention, verbal learning, verbal memory, semantic fluency, working memory and cognitive control*. The cognitive tests were from the MATRICS Consensus Cognitive Battery (MCCB) (Mohn et al., 2012; Nuechterlein et al., 2008), Halstead-Reitan (Klove, 1963), the Wechsler Adult Intelligence Scale (WAIS-III) (Wechsler, 1997), Delis Kaplan Executive Functioning System (D-KEFS) (Delis et al., 2001), and also included California Verbal Learning Test (CVLT-II) (Delis et al., 1987), and the Hopkins Verbal Learning Test-Revised (HVLT-R) (Benedict et al., 1998). Intellectual functioning was assessed using the Matrix Reasoning and Vocabulary subtests from the Wechsler Abbreviated Scale of Intelligence (WASI) (Wechsler, 2011). Case-control studies of performance on these tests have previously been published by our group (Flaaten et al., 2023, 2022; Haatveit et al., 2021; Laskemoen et al., 2020; Lunding et al., 2023; Simonsen et al., 2010), consistently showing group-level differences with more pronounced impairments in SZ, followed by BD, compared to HC. Overview and comparison between scores on similar tests from the two batteries can be found in Supp. Table 3 and Fig. S1.

### Peripheral inflammatory-immune markers

Blood was sampled from the antecubital vein in EDTA vials, stored at 4°C overnight, before isolation of plasma that was stored at −80 °C. Average freezer storage time was 5 years and freezer time was included as covariate. The peripheral inflammatory-immune markers and mediatory (descriptive table in Supp. Table 4) included: High sensitivity CRP (hsCRP), soluble tumor necrosis factor receptor 1 (sTNFR1), Interleukin-1 receptor antagonist (IL-1Ra), chitinase-3-like protein 1 (YKL-40), myeloperoxidase (MPO), Von Willebrand factor (vWF), cathepsin S (CatS), insulin-like growth factor-binding protein 4 (IGFBP4), soluble interleukin- 2 receptor (sIL-2R), chemokine ligand 16 (CXCL16), glycoprotein 130 (Gp130), CD166 antigen transmembrane glycoprotein (Alcam), cluster of differentiation 14 (CD14), galectin 3 (Gal3), protein deglycase DJ-1 (Park7), brain-derived neurotrophic factor (BDNF), Dickkopf-related protein 1 (DKK1), osteoprotegerin (OPG), pentraxin 3 (PTX3), and macrophage migration inhibitory factor (MIF). Case-control studies on a selection of these markers have previously been published by our group (Aas et al., 2017; Dieset et al., 2019; Hjell et al., 2022; Hope et al., 2010, 2009; Hoseth et al., 2016; Mørch et al., 2019, 2017, 2016; Reponen et al., 2020; Szabo et al., 2020; Werner et al., 2022).

Plasma levels of the above markers were measured in duplicate by enzyme immunoassays (EIA) using commercially available antibodies (R&D Systems, Minneapolis, MN, USA) in a 384-format using a combination of SELMA (Jena, Germany) and a BioTek (Winooski, VT, USA) dispenser/washer. Absorption was read at 450 nm with wavelength correction set to 540 nm using an ELISA plate reader (Bio-Rad, Hercules, CA, USA). All EIA’s had intra- and inter-assay coefficients <10%. A validation of the stability of the markers regarding effects of diurnal and postprandial variation as well as data from 4 samples stored at 4°C for 24h before processing has been published previously (Ormerod et al., 2022; Reponen et al., 2020; Sæther et al., 2023).

### Generation of iPSCs and differentiation of astrocytes and NPCs

Generation and maintenance of iPSCs and differentiation of astrocytes and NPCs has previously been described in detail (Akkouh et al., 2020; Osete et al., 2021). Quality control, phenotyping, monitoring of morphology and pluripotency marker expressions was conducted at the Norwegian Core Facility for Human Pluripotent Stem Cells at the Norwegian Center for Stem Cell Research. In brief, fibroblasts isolated from SZ and HC donors were reprogrammed into iPSCs using Sendai virus, transduced with the CytoTune^TM^-IPS 2.0 Sendai Reprogramming Kit (Thermo Fisher, Waltham, MA, USA). The authenticity and normality of the iPSC lines was confirmed using karyotyping of their chromosomal integrity at passage 15 (KaryoStat Karyotyping Service, Thermo Fisher). Phenotypic and functional characterization of the iPSC-astrocyte lines has previously been published (Szabo et al., 2021). Further details on astroglia and NPC differentiation are found in Supp. Methods 1.

### Inflammatory modulation of iPSC-derived astrocytes and NPCs

To mimic chronic, low-grade, as well as acute inflammatory challenge *in vitro*, astrocytes and NPCs were treated with the pro-inflammatory cytokine IL-1β (Peprotech) at working concentrations of either 0.5 (“low/chronic dose”) or 10 ng/ml (“high/acute dose”) based on a previously established treatment protocol (Akkouh et al., 2020). IL-1β is a prototypical upstream inflammatory cytokine with an important role in neuroinflammation and cognition (Shaftel et al., 2008). Culture supernatants were collected after 24 h of incubation in case of 10 ng/ml treatment, or after 7 days of repeated low dose treatments (0.5 ng/ml IL-1β on every other day). Collected samples were analyzed by ELISA as described above. Harvested samples were immediately transferred and stored at −80 °C before analyses.

### RNA microarray analysis

We analyzed microarray data from whole blood mRNA collected in Tempus Blood RNA Tubes (Life Technologies Corp.) that was available in a subcohort of the main sample (n=776) as previously described (Akkouh et al., 2018; Sheikh et al., 2022). Briefly, 200 ng of total RNA was biotin labeled and amplified using the Illumina TotalPrep-96 RNA Amplification Kit (Thermo Fisher Scientific), and global gene expression quantification was carried out with Illumina HumanHT-12 v4 Expression BeadChip (Illumina Inc.). Multidimensional scaling and hierarchical clustering were used for quality control, as well as elimination of multiple batch effects (RNA extraction batch, RNA extraction method, DNase treatment batch, cRNA labeling batch, and chip hybridization). Computational estimation of cell type abundances was performed with the recently developed BayesPrism (Chu et al., 2022), a Bayesian method to predict cellular composition and gene expression in individual cell types from bulk RNA-seq. Outliers were pruned using detectOutliers in the R package lumi (Du et al., 2008). Expression signatures from Zhang et al. (2016) and the Human Protein Atlas (https://www.proteinatlas.org/about/download) were used as prior information to deconvolve the CMC and the blood expression data, respectively. Default parameters were used in all analyses.

### RNA-sequencing of postmortem brain samples from the CMC

We evaluated RNA-seq data of postmortem dorsolateral prefrontal cortex (dlPFC) samples from 474 donors (SZ=214, BD=45, HC=215) of Caucasian ethnicity obtained from the CMC (Hoffman et al., 2019). Details on extraction, yield, quality control and RNA-seq have been previously described in Sheikh et al. (2022). All cases had a read count of >25 million reads (mean: 39.2 million). A prefiltering step excluded all lowly expressed genes (<1 count per million in >50% of cases), retaining 16,895 genes for analysis. Differential expression analyses were carried out using the limma R package (Ritchie et al., 2015) and recommended guidelines (Law et al., 2018). Gene expression reference profiles for human astrocytes, neurons, oligodendrocytes and endothelial cells were obtained from Zhang et al. (2016) for cell type-specific expression patterns.

### Statistical procedure

#### Data preprocessing

Data preprocessing, statistical analyses and visualization of results were conducted in the R-environment (https://www.r-project.org/; v.4.2.0; R-packages reported in Supp. Methods 2). Peripheral inflammatory-immune data was pruned for outliers and replaced with NA using 1.5 x IQR below or above the 25^th^ and 75^th^ percentile, then log transformed. Missing cognitive and inflammatory-immune marker data were imputed using Multiple Imputation by Chained Equations (MICE; see Supp. Table 5 and Fig. S2). Robust one-way analyses of cognitive scores and inflammatory-immune marker levels are found in Supp. Tables 6-7. Main sample and clinical characteristics were compared across groups using Kruskal-Wallis rank sum test and permutation (*n*=10000) based t-tests for continuous variables, and chi-squared tests for categorical variables. We used a previously described analysis pipeline (Sæther et al., 2023) for canonical correlation analysis (CCA) and hierarchical clustering.

#### Canonical correlation analysis (CCA)

CCA was used to identify covariance patterns between cognitive domains and peripheral marker levels across diagnostic groups and HC. In brief, CCA generates modes of covariance between two sets of variables (canonical variates), where each mode can reflect different patterns in the data. The significance of the correlation between the variable sets was evaluated using the Wilks Lambda test statistic and permutation testing (*n*=10000), repeating the CCA in each permutation by randomly shuffling the rows of the peripheral markers. We only evaluated modes performing well in a hold-out test variable set, by applying a 10-fold cross-validation procedure with *n*=100 repetitions. In each repetition a new fold was assigned as the test set (20% of the sample) and training set (80%) which was submitted to CCA. The average correlation from the training set was calculated and applied to the out-of-sample test set. We assessed the stability of the contribution of the variables using a jack-knife procedure (Dinga et al., 2019). See Supp. Methods 3 for details. Associations between participant loading scores (i.e. mode weights) of the cognitive and peripheral marker modes and group was assessed using linear regression, adjusting for age, sex, DDD of psychopharmacological treatments (SMI only), BMI, CRP, and freezer storage time.

#### Hierarchical clustering

Hierarchical clustering was applied on the cognitive and peripheral marker mode loading scores to detect subgroups. In brief, the procedure included: 1) generating a Euclidian distance matrix between loading scores, 2) choosing a linkage method based on best performing agglomerative coefficient (average, single, complete, Ward’s), and 3) determining the optimal number of clusters by inspecting the average silhouette index. The presence of clusters was tested using a previously described simulation procedure from Dinga and colleagues (Dinga et al., 2019), and the stability of the clusters was assessed using a resampling procedure (bootstrapping). The Jaccard similarity index for clustering stability was then computed (0-100%), where an index >0.7 was considered stable. See Supp. Methods 4 for further details. Cluster comparisons on the variables identified by the CCA and demographic characteristics were performed using Welch’s t-test (Bonferroni corrected).

#### Assessing the effect of inflammatory modulation on iPSC-derived astrocytes and NPCs

Candidate markers sharing variance with cognitive domains (as identified by CCA) were measured using ELISA following IL-1β treatment of astrocytes and NPCs. Differences in cytokine secretion between conditions (high/low/no treatment), and donor group (SZ vs. HC), as well as differences in cognitive domains between donors were assessed using Bayesian t-tests and ANOVAs. We followed the typical convention for interpreting Bayes Factor (BF), with BF<0.33 as substantial evidence for the null hypothesis (H_0_), BF=0.33-1 as anecdotal evidence for the H_0_, BF=1-3 as anecdotal evidence for the alternative hypothesis (H_1_), and BF>3 as substantial evidence for the H_1_ (Keysers et al., 2020). Variance was assessed using Levene’s test. We additionally assessed correlations (Pearson’s) between cytokine secretion and cognitive performance of donors.

#### Analyses of RNA microarray and RNA-seq of postmortem brain samples

Regulation of candidate markers sharing variance with cognitive domains was evaluated in leukocytes in a subcohort of our main sample, using linear regression with age, sex, BMI, DDD and ARNTL expression (i.e. to adjust for differences in time of blood sampling and circadian rhythm) as covariates (Ko and Takahashi, 2006). This was evaluated across cluster membership identified by hierarchical clustering of the covariance patterns in the main sample, and across diagnostic categories. For the CMC data, we defined high-low inflammation subgroups based on IL-6, IL-8, IL-1β and SERPINA3, previously established for subgroup classification in postmortem brain tissue in SMI (Cai et al., 2020; Fillman et al., 2013). Individuals with SMI and HC with > median expression on a composite of these markers were classified as belonging to a high inflammation group, the rest to a low inflammation group. FDR corrected linear regressions of the association between high-low inflammation subgroups, as well as across diagnostic categories, and cell type-specific RNA levels of candidate marker genes and relevant receptors was assessed with age, sex, postmortem interval, institution (laboratory batch effects) as covariates.

### Code availability

Main analysis code/scripts are available at: https://osf.io/32xrk/

## Results

### Sample demographics

Sample demographics are provided in Table 1.

**Table 1.** Sample demographics.

| Characteristics | SZ<br>(N = 435)<sup>1</sup> | BD<br>(N = 218)<sup>1</sup> | HC<br>(N = 582)<sup>1</sup> | *p*-value<sup>2</sup> | Group<br>comparisons<sup>3</sup> |
| --- | --- | --- | --- | --- | --- |
| Age | 30.0 (9.67) | 34.6 (12.3) | 33.0 (9.4) | <0.001 | SZ < HC, BD |
| Sex (female) | 174 (40%) | 131 (60%) | 273 (47%) | <0.001 | SZ, HC ~ BD |
| Education (years) | 12.4 (2.5) | 13.3 (2.5) | 14.3 (2.3) | <0.001 | SZ < BD < HC |
| WASI IQ (2-subtests) | 101 (15) | 108 (12) | 112 (11) | <0.001 | SZ < BD < HC |
| BMI (kg/m<sup>2</sup>) | 26.3 (5.2) | 25.9 (4.5) | 24.6 (3.6) | <0.001 | SZ, BD > HC |
| CRP | 3.8 (3.7) | 4.0 (4.5) | 2.8 (3.0) | <0.001 | SZ, BD > HC |
| PANSS Negative | 13.4 (6.0) | 8.5 (3.56) |  | <0.001 | SZ > BD |
| PANSS Positive | 9.9 (4.12) | 6.0 (2.9) |  | <0.001 | SZ > BD |
| PANSS Disorganized | 5.5 (2.5) | 4.2 (1.7) |  | <0.001 | SZ > BD |
| PANSS Excited | 5.6 (2.1) | 5.3 (1.8) |  | ns | - |
| PANSS Depressed | 8.3 (3.23) | 7.6 (2.9) |  | <0.05 | SZ > BD |
| YMRS | 5.1 (5.1) | 3.8 (5.1) |  | <0.001 | SZ > BD |
| GAF Symptom | 42.7 (11.6) | 57.0 (11.9) |  | <0.001 | SZ < BD |
| GAF Function | 44.1 (11.4) | 53.1 (12.7) |  | <0.001 | SZ < BD |
| Age at onset | 23.7 (8.4) | 26.8 (10.5) |  | 0.003 | SZ < BD |
| Duration of illness (years) | 6.4 (7.3) | 7.7 (9.1) |  | ns | SZ < BD |
| Antipsychotics, DDD | 1.1 (1.01) | 0.4 (0.65) |  | <0.001 | SZ > BD |
| Antidepressants, DDD | 0.5 (0.92) | 0.4 (0.74) |  | ns | - |
| Antiepileptics, DDD | 0.0 (0.26) | 0.3 (0.52) |  | <0.001 | SZ < BD |
| Lithium, DDD | 0.0 (0.14) | 0.2 (0.49) |  | <0.001 | SZ < BD |
| Total, DDD | 1.7 (1.48) | 1.4 (1.17) |  | <0.05 | SZ > BD |

<sup>1</sup>Mean (SD); n (%)
<sup>2</sup>Kruskal-Wallis rank sum test; Pearson's Chi-squared test
<sup>3</sup>Pairwise two-sample Permutation Test (for 3 groups)
Abbreviations: HC, healthy controls; SZ, schizophrenia; BD, bipolar disorder; WASI, Wechsler Abbreviated Scale of Intelligence; BMI, body mass index; CRP, C-reactive Protein; PANSS, Positive and Negative Syndrome Scale; YMRS, Young Mania Rating Scale; GAF, Global Assessment of Functioning scale; DDD, defined daily dosage

### Multivariate pattern of cognitive functioning and peripheral markers (CCA)

The CCA revealed one significant covariance mode explaining 17% of the variance (r=0.41) capturing psychomotor processing speed, fine motor speed and verbal learning, which correlated with a combination of markers reflecting vascular and endothelial-related inflammation, innate immune activation, and ECM remodelling, including YKL-40, hsCRP, CatS, sTNFR1 and MIF (Fig. 3 A-C). Decreasing loading scores indicate a pattern of lower cognitive functioning and higher peripheral marker levels, whereas an increase in loading scores indicate higher cognitive functioning and lower marker levels. Canonical loadings showed stability (Fig. S4). Detailed statistical results of the CCA are found in Supp. Results 1. Both SZ (loading score estimate=-0.97±0.07, *t*=-14.59, *p*<0.001), and BD (loading score estimate=-0.41±0.07, *t*=-5.44, *p*<0.001) was associated with lower loading scores for the cognitive canonical variate relative to HC. The same pattern was observed for the inflammatory-immune canonical variate, for SZ (loading score estimate=-0.48±0.08, *t*=-6.03, *p*<0.001) and BD (loading score estimate=-0.35±0.09, *t*=-3.96, *p*<0.001). See Fig. 2 D-E for loading score differences on both canonical variates between SZ, BD, and HC groups.

**Figure 2.**
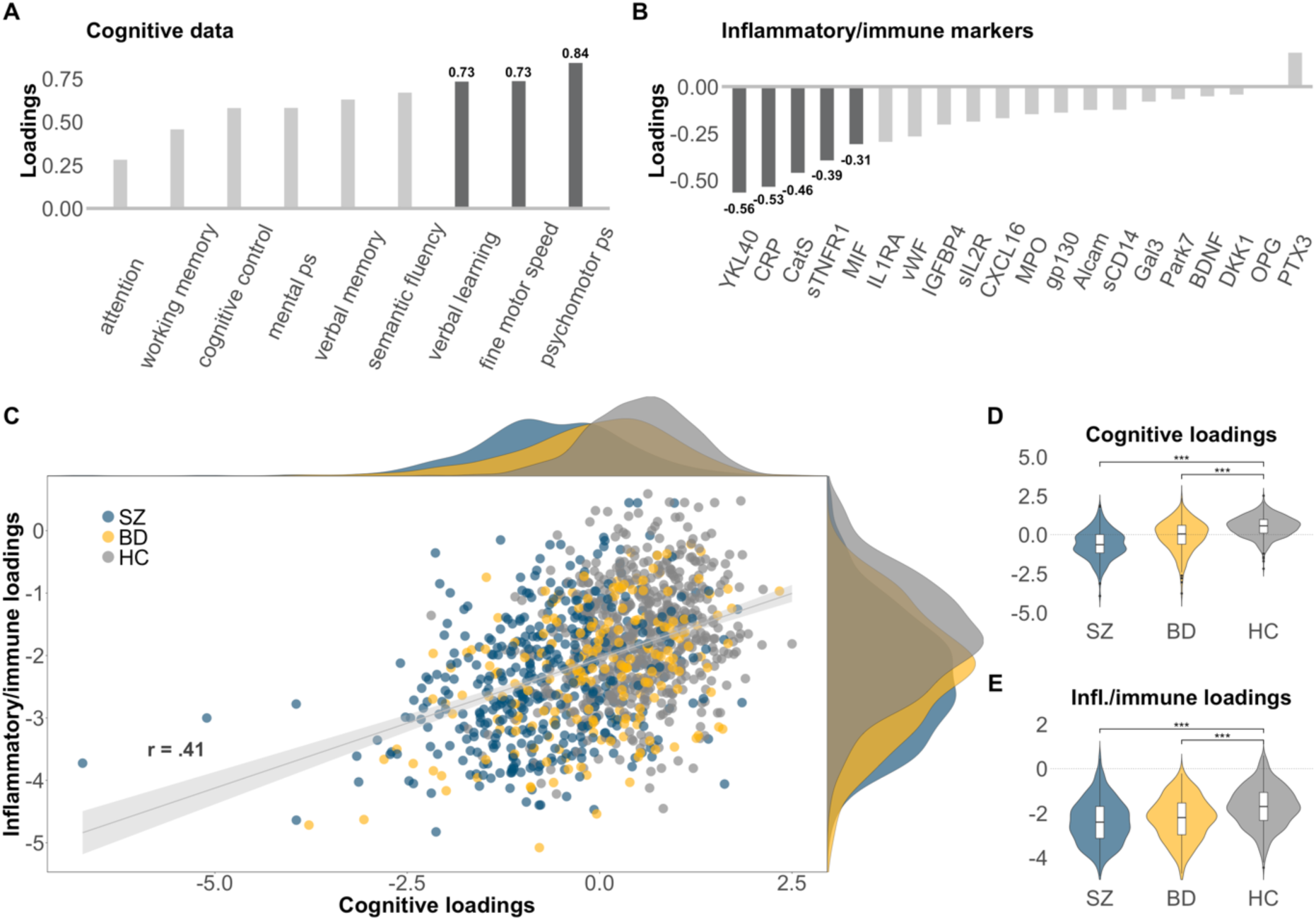
Covariance pattern between cognitive and inflammatory/immune marker data in SZ, BD and HC. Panel A-B shows contributions of each variable in the cognitive and inflammatory/immune marker datasets to the significant mode (top variables in bold). Panel C shows the correlation between the individual loading scores, including distribution density (top = cognitive loading scores, right = inflammatory/immune marker loading scores). Panel D-E shows differences the loading scores between SZ, BD and HC. (*p*<0.001=***, *p*<0.01=**, *p*<0.05=*).

**Figure 3.**
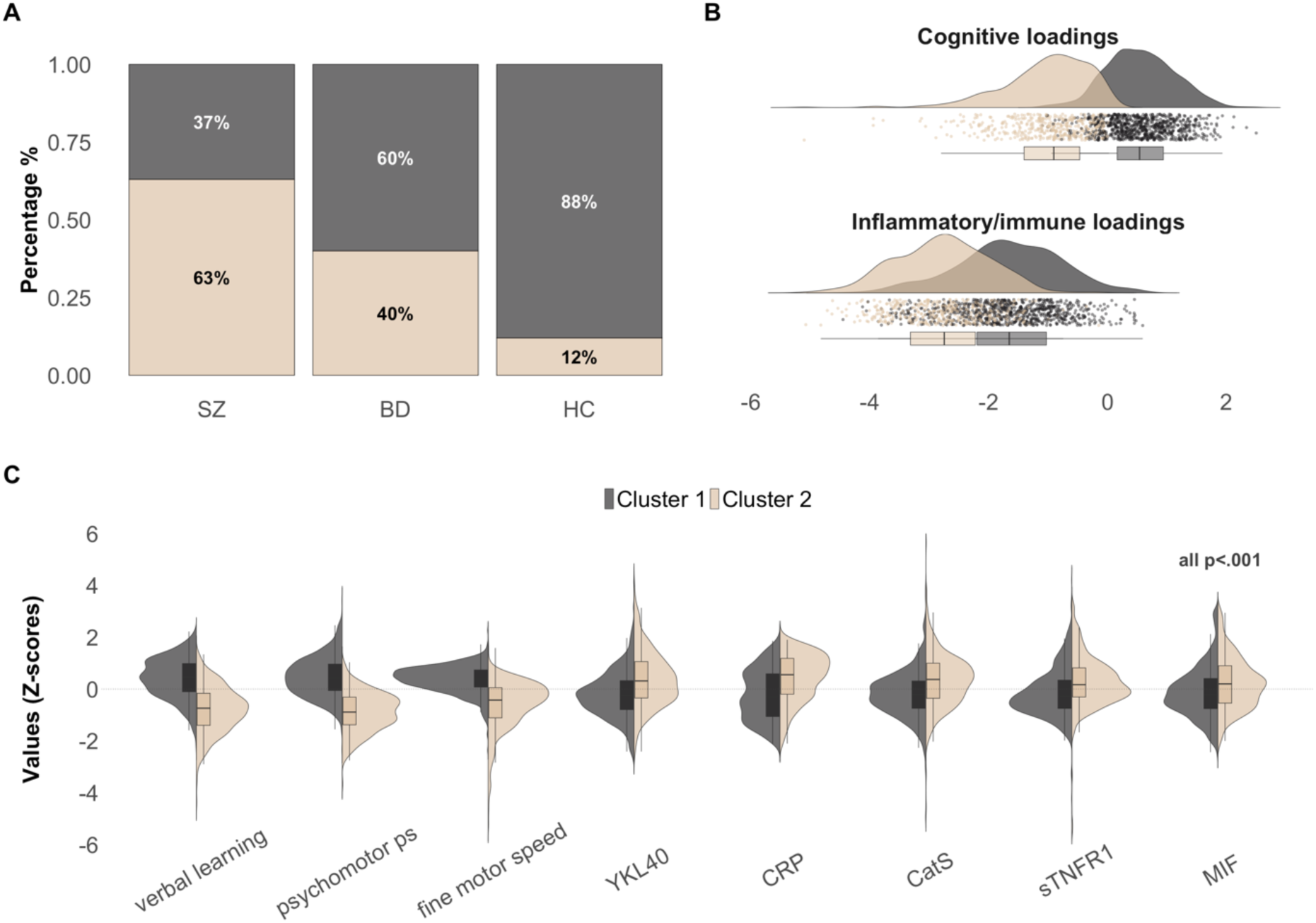
Hierarchical clustering of covariance pattern. Panel A shows the percentage of individuals with SZ, BD and HC assigned to each cluster. Panel B shows the difference in loading scores between the clusters. Panel C shows cluster differences across variables identified in the CCA.

### Subgroups of cognitive and immune-inflammatory patterns

Hierarchical clustering and related analyses revealed a 2-cluster solution to be optimal (Supp. Results 2). The first cluster captured a subgroup exhibiting more positive loading scores on both canonical variates (*n*=807, SZ=161 [37%], BD=131 [60%], HC=515 [88%]), whereas the second subgroup (*n*=428, SZ=274 [63%], BD=87 [40%], HC=67 [12%]) was characterized by more negative loading scores (all *p*<0.001, see Fig. 3 A-C). Follow-up analyses confirmed that the second subgroup (cluster 2) had higher levels of the peripheral inflammatory-immune markers and lower scores on the cognitive domains identified by the CCA, compared to the first subgroup (all *p*<0.001). Individuals in the second subgroup were characterized by lower IQ and shorter education, higher BMI, and higher age (all *p*<0.001) compared to the first subgroup. Individuals with SMI in the second subgroup had more negative, positive, and disorganised symptoms, lower functioning (GAF-S, GAF-F), and used a higher dose of antipsychotics as well as total DDD score compared to individuals in the first subgroup (all *p*<0.001). See Supp. Table 8 for details.

### Effects of inflammatory modulation on candidate marker secretion in iPSC-derived astrocytes and NPCs

We next investigated the effects of IL-1β treatment on the secretion of YKL-40, CatS, sTNFR1 and MIF, markers that were increased in peripheral blood in the second subgroup (consisting of primarily SZ), in iPSC-derived astrocytes and NPCs from SZ and HC donors. There was limited evidence of a difference between low/chronic, high/acute and no IL-1β treatment conditions, except for increased CatS secretion in NPCs (BF=82.33) and astrocytes (BF=3.53) in IL-1β treatment conditions relative to no treatment. There was also limited evidence for a difference in cytokine secretion between SZ and HC donors. However, YKL-40 was the only marker slightly favoring the H_1_ in astrocytes low (BF=1.18) and high (BF=1.39) IL-1 β treatment conditions, as well as in unstimulated astrocytes (BF=1.47) and NPCs (BF=1.59). As seen in Fig. 4A there was larger variance in YKL-40 secretion in SZ donors compared to HC donors (Levene’s test all *p*<0.01, except NPCs), suggestive of heterogeneity (additional info in Supp. Table 9-10). Finally, we investigated the relationship between YKL-40 secretion levels and candidate cognitive domain scores from SZ and HC donors. There was a negative correlation between YKL-40 secretion in iPSC-derived astrocytes and fine motor speed (r=-0.78, *p*=0.013) in the low IL-1 β treatment condition (Fig. 4B).

**Figure 4.**
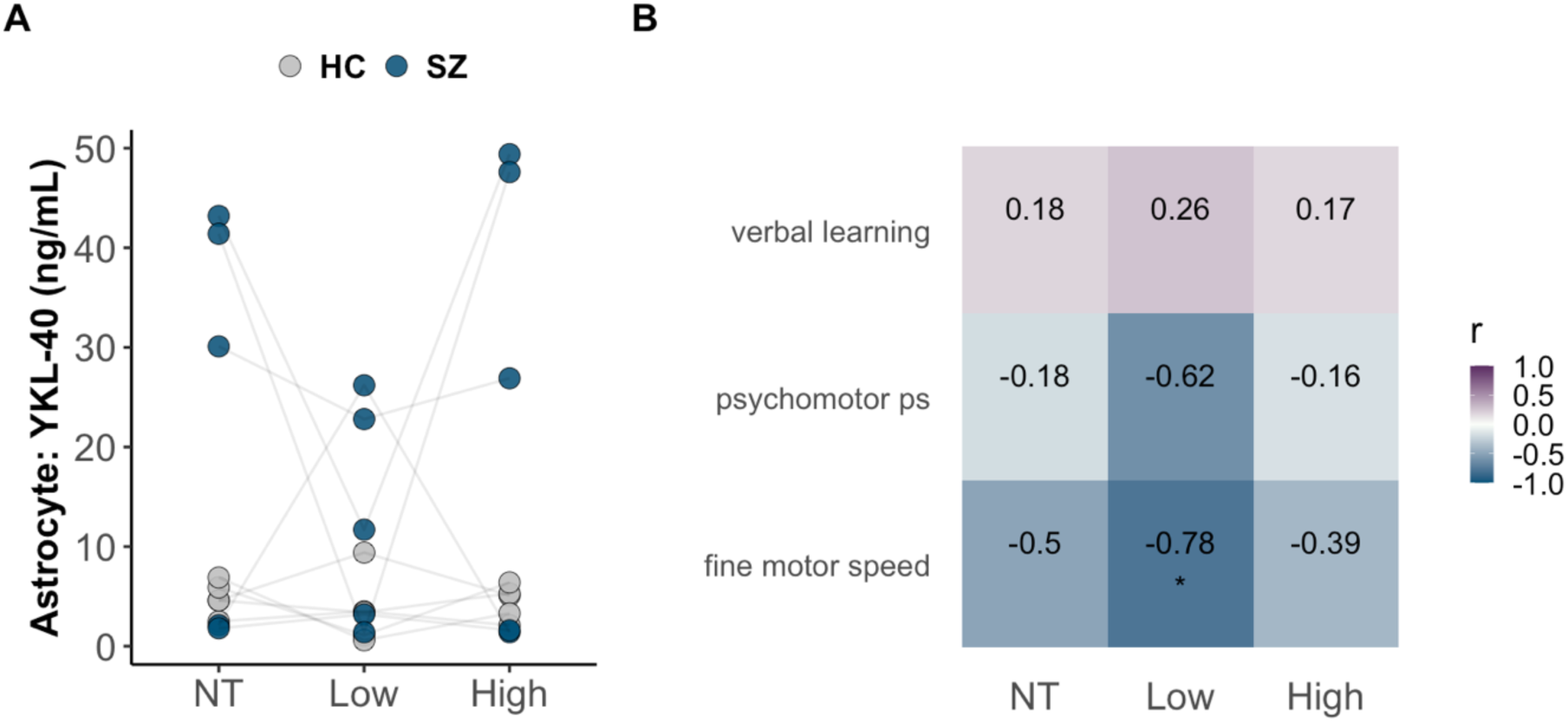
In vitro results and comparison with donor cognitive scores. Panel A shows the difference in YKL-40 secretion following no treatment, low and high IL-1β concentration between iPSC-derived astrocytes from SZ and HC donors. Panel B shows the correlation matrix between YKL-40 secretion levels in astrocytes and candidate cognitive domain scores from SZ and HC donors, with fine motor speed significant after Bonferroni correction(*p*=0.013).

### Evaluation of YKL-40 and related gene expression in circulating leukocytes and postmortem brain samples

Analyses so far suggested that YKL-40 could be of importance for mediating effects on cognition in SMI as assessed by large-scale analyses in peripheral blood in the main sample and results from the iPSC experiments. To evaluate if circulating leukocytes could be a source of increased peripheral levels of the candidate markers, we evaluated their mRNA expression in a subcohort of the main sample. We found no evidence of dysregulated expression of *CHI3L1* (encoding YKL-40), *CTSS* (encoding CatS), *MIF* and *TNFRSF1A* (encoding TNFR1) in isolated leukocytes from whole blood between clusters or diagnostic groups (Fig. 5A), suggesting other sources.

**Figure 5.**
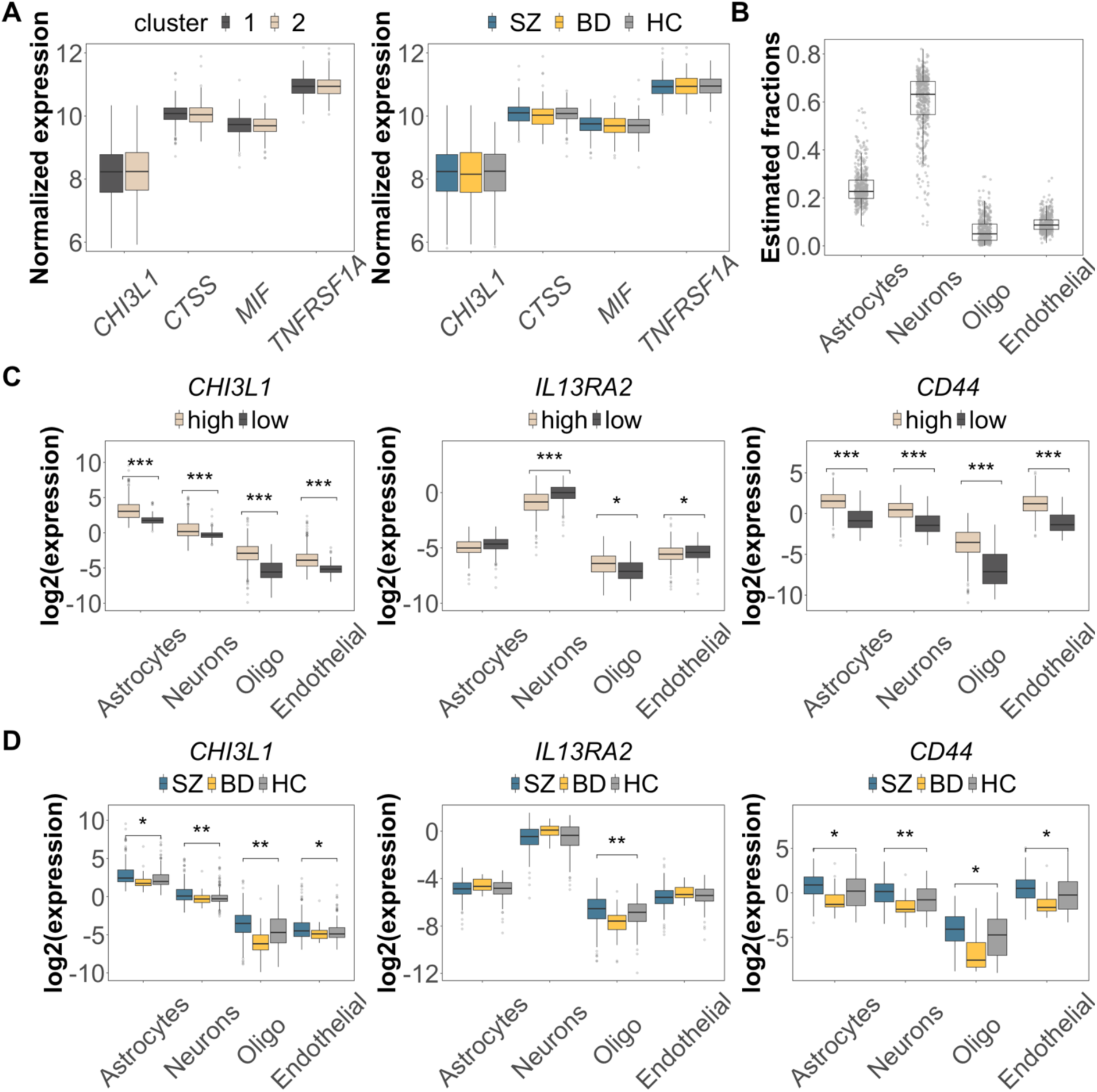
YKL-40 and related gene expression in leukocytes and postmortem brain samples. Panel A shows similar expression of candidate genes in leukocytes across both clusters and across diagnostic groups. Panel B shows estimated fractions of brain cells following computational deconvolution. Panel C shows cell type-specific expression of *CHI3L1*, *IL13RA2* and *CD44* in DLPFC samples from the CMC, between high and low inflammation groups. Panel D shows the same as C but between diagnostic groups. (*p*<0.001=***, *p*<0.01=**, *p*<0.05=*, FDR corrected).

To evaluate the relevance of our *in vitro* finding of somewhat higher, but heterogeneous, YKL-40 secretion in astrocytes in SZ, we evaluated the *in vivo* expression of *CHI3L1* and putative receptors *IL13RA2* and *CD44* in brain samples obtained from the CMC. Estimated fractions of brain cells following computational deconvolution was as expected (Fig. 5B). The markers used to define high-low inflammatory subgroups showed positive correlation with the expression of candidate markers, except for IL-8 (Fig. S7). The high inflammation group consisted predominantly of SZ (SZ=124, BD=14, HC=99, chi square *p*<0.01), and the low inflammation group consisted primarily of HC (SZ=90, BD=31, HC=116, chi square *p*<0.01). As shown in Fig. 5C, expression levels were generally higher across brain cells for the high inflammation group, except for neuronal levels of *IL13RA2*. Notably, even within the high and low inflammation groups, SZ had significantly higher *CHI3L1* expression compared to HC (*p*<0.05, 12% and 9% higher in SZ, respectively), but there was no difference between BD and HC. Evaluation in relation to diagnostic categories revealed higher levels of CHI3L1 and CD44 across all brain cells in SZ compared to HC, while BD displayed similar levels to HC, presumably due to low BD sample size (Fig. 5D).

## Discussion

This study aimed to explore a potential link between peripheral and central inflammatory-immune activity and cognitive impairment in SMI. First, we identified transdiagnostic subgroups based on covariance between cognitive domains (measures of psychomotor processing speed, fine-motor speed, and verbal learning) and peripheral markers reflecting inflammatory response (CRP, sTNFR1, YKL-40), innate immune activation (MIF) and ECM remodelling (YKL-40, CatS). Of the candidate markers, we found evidence of increased variance in YKL-40 secretion in iPSC-derived brain cells in SZ compared to HC donors. Notably, *CHI3L1*, the gene encoding for YKL-40 and *CD44*, known to interact with YKL-40, was higher in postmortem brain samples across all types of brain cells in SZ and in a high inflammatory subgroup consisting of a larger proportion of SZ. Our findings confirm that immune-cognition associations are heterogeneous and transdiagnostic, and highlight a potential pathophysiological role for YKL-40 in cognitive impairment in SMI.

Based on covariance patterns in our main sample we identified a more compromised low cognition – high inflammatory-immune dysregulation subgroup, consisting primarily of individuals with SMI. This is a consistent pattern also observed in our previous work using a different inflammatory-immune marker panel (Sæther et al., 2023), and is in line with evidence that individuals with SMI in high inflammatory subgroups tend to have lower cognitive functioning (Fillman et al., 2016; Lizano et al., 2023a, 2020). We also found that individuals with SMI with co-occurring inflammation and cognitive impairment have more symptoms and lower functioning, which may be important to consider in future clinical trials. Interestingly, we found the same cognitive domains as in our previous work (Sæther et al., 2023), i.e. measures of speed and verbal learning, shared variance with inflammatory-immune markers. Inflammation has long been associated with psychomotor slowing across SMI (Felger and Treadway, 2017; Goldsmith et al., 2016; Goldsmith et al., 2020; Larsen et al., 2021), and in experimentally induced inflammation (Brydon et al., 2008; Reichenberg et al., 2001). Reduced psychomotor processing speed and verbal learning performance have recently been linked to altered brain functional connectivity in high inflammation subgroups in SMI (Lizano et al., 2023a). This could indicate that these cognitive domains are particularly relevant in the context of inflammatory-immune dysregulation in SMI.

Of the candidate peripheral markers, YKL-40 stood out in *in vitro* and *in vivo* analyses. Similar to our findings from plasma, we also observed heterogeneity in the secretion of YKL-40 in iPSC-derived astrocytes. The majority of SZ donors exhibited high YKL-40 secretion while others had similar levels to HC, regardless of inflammatory challenge. Indeed, peripheral levels of YKL-40 is associated with genetic variation in the CHI3L1 gene (Nielsen et al., 2011; Ridker et al., 2014). In population studies, a SNP in the promoter region have been associated with both risk of SZ and higher peripheral levels of the cytokine (Yamamori et al., 2012; Zhao et al., 2007). In the postmortem brain samples, individuals belonging to the high inflammatory subgroup showed higher expression of particularly *CHI3L1* and *CD44*, known to interact (Zhao et al., 2020), which was not specific to cell type. Individuals with SZ showed a more apparent dysregulation pattern, however, more BD data is needed to determine whether there are distinct mechanisms between diagnostic groups. Since no cognitive data exist from individuals in the CMC cohort, it is not possible to determine a presence of co-occurring psychomotor slowing in the high inflammation subgroup. However, we did find a negative correlation with donor performance on psychomotor processing speed. While caution is needed when interpreting small sample *in vitro* studies, the results warrant further investigation of a potential role of YKL-40 in psychomotor slowing in SMI.

Produced and secreted by various CNS cells, YKL-40 has pleiotropic functions including tissue remodelling, pathogen defense and reactive gliosis (Pinteac et al., 2021; Ziatabar et al., 2018). Several studies have shown dysregulated levels of YKL-40 in SMI (Arion et al., 2007; Dieset et al., 2019; Horváth and Mirnics, 2014; Jakobsson et al., 2015; Rolstad et al., 2015; Zhao et al., 2007). It is being considered as a biomarker of disease progression and cognitive impairment in cardiovascular-, neurodegenerative-, autoimmune- and inflammatory diseases (Kjaergaard et al., 2016; Li et al., 2023; Moreno-Rodriguez et al., 2020; Talaat et al., 2023; Tizaoui et al., 2022; Yeo et al., 2019). While its exact mechanisms of action are still unknown, it is speculated that accumulation of chitinase-like proteins are toxic to CNS cells and lead to astrogliosis, potentially impacting cognitive functioning (Li et al., 2023; Lomiguen et al., 2018; Turano et al., 2015). Indeed, increased frontal cortex expression of YKL-40 in neurodegenerative disease has been associated with poorer episodic memory and perceptual speed (Llorens et al., 2017; Moreno-Rodriguez et al., 2020). YKL-40 has also been suggested as a glial marker of inflammation-induced synaptic degeneration in Alzheimer’s and related dementias (Hellwig et al., 2015). Its upregulation is also observed in response to inflammatory stimuli including IL-6, interferon-γ, and TNFα, all linked to SMI (Goldsmith et al., 2016; Halstead et al., 2023; Upthegrove et al., 2014).

The two putative YKL-40 receptors we assessed have mostly been studied in brain tissue in the context of glioblastomas (Guetta-Terrier et al., 2023; Wu and Low, 2003). Interestingly, YKL-40, via IL13RA2, is also involved in regulating inflammasome activation (Cruz et al., 2012; He et al., 2013), which we have previously shown to be dysregulated in SMI (Szabo et al., 2022), and related to cognitive impairment (Sæther et al., 2022). However, of these two receptors, CD44 displayed a pattern similar to YKL-40, with a higher expression in the high-inflammation subgroup and in individuals with SMI. Its regulation has been shown to affect cognition in experimental models (Raber et al., 2014; Sun et al., 2020), and may activate Erk and Akt pathways and Wnt signalling (Geng et al., 2018), pathways associated with cognitive impairment (Albert-Gascó et al., 2020; Oliva et al., 2013; Shu et al., 2013). Of relevance, activation of the adenosine A2a receptor (ADORA2A), may decrease YKL-40 expression in astrocytes and alleviate neuroinflammation and white matter injury (Yuan et al., 2022). Dysregulation of ADORA2A in astrocytes has been shown to disrupt glutamate homeostasis leading to cognitive impairment, and in a SZ subgroup, reduced striatal ADORA2A levels is accompanied by an altered psychomotor phenotype (Matos et al., 2015). Given our findings and general evidence of a role of YKL-40 across brain disorders, accompanied by a corresponding upregulation of CD44, future studies should evaluate if and how YKL-40 could promote neuroinflammation and cognitive impairment in SMI. Should that be the case, the next step will be to consider if this marker represents a therapeutic target with clinical utility.

There are some limitations to consider. The cross-sectional design does not allow for inference regarding a potential causal or bidirectional relationship between inflammatory-immune activity and cognitive functioning, which may be influenced by numerous factors not accounted for in this study. While our study can elucidate potentially relevant immune-related mechanisms underlying cognitive impairment, we still need longitudinal data and replication in independent samples to determine the clinical relevance. Our findings do not exclude the potential importance of other inflammatory-immune markers and mediators, and several pathophysiological processes are likely involved in the cognitive impairments observed in SMI (McCutcheon et al., 2023). We included storage duration as a covariate in our models due to potential protein degradation, which could vary from protein to protein. However, we have previously determined a high degree of correlation (*r*=0.86) between CRP during isolation of plasma, and CRP determined years later during bulk analysis in an overlapping sample (Reponen et al., 2020). Further, the donors selected for iPSC-derived cell lines were not based on prior cognitive information, and future studies should consider selecting donors based on cognitive phenotypes. Finally, human iPSC studies typically have low sample sizes due to costs, long differentiation protocols and other technical challenges. However, the heterogeneity observed in our study and others (Lizano et al., 2023b), emphasize that increasing sample sizes will be important to improve our understanding of inflammatory-immune dysregulation underlying cognitive impairment in affected individuals with SMI.

## Conclusions

Our findings linking peripheral and central inflammatory-immune activity confirm transdiagnostic subgroups with reduced cognitive functioning and inflammatory-immune abnormalities in SMI, and point to a potential pathophysiological role of YKL-40.

## Supporting information

Supp_Res

## Data Availability

Data may be available upon reasonable request to the authors

## Acknowledgements

We wish to thank all participants for their contribution. Funding was provided by the South-Eastern Norway Regional Health Authority (grant number 2020089) and Research Council of Norway (#223273). The CMC postmortem brain sample data was obtained from NIMH Repository & Genomics Resource, a centralized national biorepository for genetic studies of psychiatric disorders (https://www.nimhgenetics.org/resources/acknowledgements). We wish to acknowledge Sigma2 (the National Infrastructure for High Performance Computing and Data Storage in Norway), and Services for Sensitive Data (TSD) at the University of Oslo.

