## Supplementary material for "Cognitive and inflammatory heterogeneity in severe mental illness: Translating findings from blood to brain": Supp_Tables

**Supplementary Tables**

| **Supplementary Table 1.** Demographic characteristics of fibroblast donors | | | | | | | | | | |
| --- | --- | --- | --- | --- | --- | --- | --- | --- | --- | --- |
|  | **SZ**  **(N = 5)** | | | | | **HC (N = 5)** | | | | |
|  | **Donor 1** | **Donor 2** | **Donor 3** | **Donor 4** | **Donor 5** | **Donor 1** | **Donor 2** | **Donor 3** | **Donor 4** | **Donor 5** |
| **Age** | In their 50s | In their 40s | In their 20s | In their 20s | In their 20s | In their 50s | In their 20s | In their 40s | In their 40s | In their 20s |
| **Sex** | male | female | male | female | female | female | male | female | female | male |
| **Ethnicity** | Caucasian | Caucasian | Caucasian | Caucasian | Mixed | Caucasian | Caucasian | Caucasian | Caucasian | Caucasian |
| Abbreviations: schizophrenia (SZ), healthy controls (HC) | | | | | | | | | | |

| **Supplementary Table 2.** Somatic medication use by SMI group | | |
| --- | --- | --- |
| **Somatic medications** | **SZ**  **(N = 435)** | **BD (N = 218)** |
| Anti-inflammatory/immunomodulatory, N (%) | 7 (1.6) | 3 (1.4) |
| Antidiabetics, N (%) | 7 (1.6) | 1 (0.5) |
| Cardiovascular/lipid modifying, N (%) | 19 (4.4) | 11 (5.0) |
| Antihistamines, N (%) | 18 (4.1) | 10 (4.6) |
| Gastrointestinal agents, N (%) | 14 (3.2) | 2 (0.9) |
| *Other, N (%) | 18 (4.1) | 44 (20.2) |
| *** Other includes vitamins, minerals, analgetics, thyroid agents, pulmonary agents, urological agents, musculoskeletal agents, contraceptives, sex hormones, anxiolytics, anti-inflammatory agents (local administrative agents, i.e. ointments/inhalators), hematological agents, parenteral nutrition agents, substance dependency agents and mucolytic agents.  Abbreviations: schizophrenia (SZ), bipolar disorder (BD), healthy controls (HC) | | |

| **Supplementary Table 3.** Overview of cognitive domains and corresponding tests from test battery I and II | | | | |
| --- | --- | --- | --- | --- |
| ***Domain*/Test^a^** | **SZ**  Battery I (N=359)  Battery II (N=76) | **BD**  Battery I (N=198)  Battery II (N=20) | **HC** Battery I (N=378)  Battery II (N=204) | **Test battery** |
| *Psychomotor processing speed* |  |  |  |  |
| Symbol Coding (WAIS-III) | -0.5 (0.9) | -0.1 (0.9) | 0.5 (0.8) | I |
| BACS Symbol Coding (MCCB) | -0.6 (1.0) | -0.6 (0.8) | 0.3 (0.9) | II |
| *Verbal learning* |  |  |  |  |
| Total recall (CVLT-II) | -0.4 (1.0) | 0.2 (1.0) | 0.3 (0.8) | I |
| Total recall (HVLT-R, MCCB) | -0.6 (1.1) | -0.2 (1.0) | 0.3 (0.8) | II |
| *Verbal memory* |  |  |  |  |
| Long-delay free recall (CVLT-II) | -0.4 (1.0) | 0.1 (1.0) | 0.3 (0.8) | I |
| Delayed recall (HVLT-R, MCCB) | -0.7 (1.2) | -0.2 (1.1) | 0.3 (0.8) | II |
| *Semantic fluency* |  |  |  |  |
| Category fluency (D-KEFS) | -0.6 (0.9) | 0.0 (1.0) | 0.5 (0.8) | I |
| Category fluency (MCCB) | -0.6 (1.0) | -0.4 (0.6) | 0.3 (0.9) | II |
| *Working memory* |  |  |  |  |
| Letter Number Sequencing (WAIS-III) | -0.4 (0.9) | -0.2 (0.8) | 0.4 (0.9) | I |
| Letter Number Sequencing (MCCB) | -0.4 (0.9) | -0.5 (0.9) | 0.2 (1.0) | II |
| *Fine motor speed* |  |  |  |  |
| Grooved Pegboard test (Halstead-Reitan) | -0.4 (1.1) | -0.2 (1.0) | 0.4 (0.5) | I & II |
| *Attention* |  |  |  |  |
| Digit Span Forward (WAIS-III) | -0.2 (1.0) | -0.1 (1.0) | 0.2 (1.0) | I & II |
| *Mental processing speed* |  |  |  |  |
| Color naming+Color reading (D-KEFS) | -0.4 (1.0) | -0.1 (1.0) | 0.3 (0.6) | I & II |
| *Cognitive control* |  |  |  |  |
| Inhibition+Inhibition switching (F-KEFS) | -0.4 (1.0) | -0.1 (1.0) | 0.3 (0.6) | I & II |
| ^a^Mean (standard deviation, SD) of Z-scores per test *Note:* Battery I N = 935; Battery II N = 300. We standardized the tests separately before collapsing in order to increase N as two different cognitive test batteries have been used. The domains are named based on the function the tests measure. Tests measuring cognitive control, attention, mental processing speed and fine motor speed were the same for both cognitive batteries. See Supplementary Figure X for visual representation stratified by group. Abbreviations: schizophrenia (SZ), bipolar disorder (BD), healthy controls (HC) | | | | |

**Supplementary Table 4. Abbreviations and descriptions of analyzed inflammatory-immune markers**

| **Abbreviation** | **Brief description** |
| --- | --- |
| hs-CRP | High-sensitivity C-Reactive Protein, general down-stream marker of inflammation |
| sTNFR1 | Soluble tumor necrosis factor receptor 1, binds to tumor necrosis factor-alpha. Can activate NF-Kb, mediates apoptosis, regulates inflammation. Induced by, and may reflect activation of TNF |
| IL-1Ra | Interleukin-1 receptor antagonist. Induced by, and may reflect activation of IL-1β |
| YKL-40 | Chitinase-3-like protein 1, inflammatory marker, tissue injury, extracellular matrix/tissue remodeling |
| MPO | Myeloperoxidase, key element of innate immune system, expressed primarily by neutrophils in defense against pathogens. Systemic levels may reflect neutrophil activation and oxidative stress |
| vWF | Von Willebrand factor, glycoprotein important for hemostasis and activation of endothelial cells |
| CatS | Cathepsin S, a lysosomal protease, plays a role in the regulation of inflammation by processing cytokines and host defense proteins, autophagy, extracellular matrix remodeling |
| IGFBP4 | Insulin-like growth factor-binding protein 4, vascular apoptosis |
| sIL-2R | Interleukin-2 receptor, expressed on the surface of immune cells. Systemic levels reflect activation of T cells |
| CXCL16 | Chemokine ligand 16, transmembrane adhesion molecule. Involved in atherosclerosis and systemic levels reflect vascular inflammation |
| Gp130 | Glycoprotein 130, member of the IL-6 family |
| Alcam | CD166 antigen, transmembrane glycoprotein. Expressed on activated T cells, monocytes, epithelial cells, fibroblasts, neurons. Mediates lymphocyte migration across central nervous system barriers |
| sCD14 | Cluster of differentiation 14, marker of monocyte activation |
| Gal3 | Galectin 3, involved in cell adhesion, cell growth and differentiation, cell cycle and apoptosis. Systemic levels may reflect fibrosis and extracellular matrix remodeling |
| Park7 | Protein deglycase DJ-1 (Parkinson disease protein 7), sensor for oxidative stress |
| BDNF | Brain-derived neurotrophic factor, found in CNS and periphery, support survival, growth and differentiation of neurons and synapses |
| DKK1 | Dickkopf-related protein 1, soluble antagonist of canonical Wnt signaling |
| OPG | Osteoprotegerin, soluble decoy receptor in TNF family. Marker of vascular inflammation |
| PTX3 | Pentraxin 3, long pentraxin in same family as CRP produced by vascular cells and may reflect local inflammation |
| MIF | Macrophage migration inhibitory factor, inflammatory cytokine in IL-6 superfamily, regulator of innate immunity |

| **Supplementary Table 5.** Percentage missing per inflammatory/immune marker and cognitive domain | | | | |
| --- | --- | --- | --- | --- |
| **Inflammatory/immune markers** | **SZ (N = 435)** | **BD (N = 218)** | **HC (N = 582)** | **Total (N = 1235)** |
|  | **Percentage (%) missing** | | | |
| sTNFR1 | 0.2 | 0 | 0 | 0.08 |
| CXCL16 | 0.9 | 0 | 0.2 | 0.4 |
| OPG | 0 | 0 | 0 | 0 |
| IL-1RA | 0 | 0 | 0 | 0 |
| sIL-2R | 0 | 0 | 0 | 0 |
| Alcam | 0 | 0.4 | 0 | 0.08 |
| gp130 | 0 | 0.4 | 0 | 0.08 |
| MPO | 0 | 0.4 | 0 | 0.08 |
| YKL-40 | 0 | 0.4 | 0 | 0.08 |
| vWF | 0 | 0.4 | 0 | 0.08 |
| BDNF | 0 | 0.4 | 0 | 0.08 |
| CatS | 0 | 0.4 | 0 | 0.08 |
| Park7 | 0 | 0 | 0 | 0 |
| PTX3 | 0 | 0 | 0 | 0 |
| IFGBP4 | 0 | 0 | 0 | 0 |
| sCD14 | 0 | 0 | 0.2 | 0.08 |
| DKK1 | 0 | 0 | 0 | 0 |
| Gal3 | 0 | 0 | 0 | 0 |
| MIF | 0.2 | 0 | 0.2 | 0.1 |
| hs-CRP | 0 | 0 | 0.2 | 0.08 |
| **Cognitive domains** | | | | |
| Fine motor speed | 2.3 | 1.4 | 3.9 | 2.9 |
| Psychomotor processing speed | 0.4 | 0 | 0.2 | 0.2 |
| Mental processing speed | 5.4 | 0 | 0.5 | 0.2 |
| Attention | 14.5 | 0.4 | 3.6 | 6.9 |
| Verbal learning | 0 | 0 | 0 | 0 |
| Verbal memory | 0.7 | 0 | 0 | 0.2 |
| Semantic fluency | 5.5 | 0 | 0.2 | 2.0 |
| Working memory | 7.8 | 12.8 | 2.2 | 6.1 |
| Cognitive control | 0.7 | 0.4 | 3.7 | 2.1 |

*Note:* Participants missing data for > 3 cognitive domains and > 4 inflammatory/immune markers were excluded (N = 26). Imputations were performed using Multiple Imputation by Chained Equations (MICE) and no variable had > 10% missing (max 6.9%). Main analyses were tested with complete cases (N = 1035) and imputed dataset (N = 1235), and both had CCA correlation r = .41 with overlap in the variables with the highest contributions to the correlation.

| **Supplementary Table 6.** Cognitive domain scores between SZ, BD and HC | | | | | | |
| --- | --- | --- | --- | --- | --- | --- |
| **Cognitive domains^a^** | **SZ  (N = 435)** | **BD  (N = 218)** | **HC  (N = 582)** | ***p*-value^b^** | **Effect size^c^** | **Pairwise comparisons^b^** |
| Fine motor speed | -0.37 (1.12) | -0.19 (1.01) | 0.36 (0.51) | ***p*<0.001** | 0.46 | SZ<HC,BD;BD<HC |
| Psychomotor processing speed | -0.56 (0.94) | -0.13 (0.93) | 0.46 (0.82) | ***p*<0.001** | 0.53 | SZ<HC,BD;BD<HC |
| Mental processing speed | -0.41 (1.02) | -0.08 (0.97) | 0.34 (0.65) | ***p*<0.001** | 0.45 | SZ<HC,BD;BD<HC |
| Attention | -0.25 (0.99) | -0.09(1.00) | 0.19 (0.97) | ***p*<0.001** | 0.24 | SZ<HC,BD;BD<HC |
| Verbal learning | -0.47 (1.01) | 0.14 (1.03) | 0.30 (0.84) | ***p*<0.001** | 0.42 | SZ<HC,BD |
| Verbal memory | -0.42 (1.06) | 0.06 (1.01) | 0.28 (0.82) | ***p*<0.001** | 0.38 | SZ<HC,BD;BD<HC |
| Semantic fluency | -0.59 (0.93) | -0.02 (0.99) | 0.44 (0.81) | ***p*<0.001** | 0.53 | SZ<HC,BD;BD<HC |
| Working memory | -0.41 (0.92) | -0.22 (0.87) | 0.36 (0.95) | ***p*<0.001** | 0.44 | SZ<HC,BD;BD<HC |
| Cognitive control | -0.36 (1.01) | -0.09 (0.96) | 0.30 (0.58) | ***p*<0.001** | 0.41 | SZ<HC,BD;BD<HC |
| ^a^Mean (standard deviation, SD), Z-scores  ^b^Robust one-way ANOVA and Lincoln post-hoc for pairwise comparison (multiple comparison corrected; WRS2 R-package)^[1]^  ^c^Explanatory measure of effect size, ξ (0.1, 0.3, 0.5, small/medium/large; WRS2 R-package)  Abbreviations: schizophrenia (SZ), bipolar disorder (BD), healthy controls (HC) | | | | | | |

| **Supplementary Table 7.** Inflammatory/immune-related marker levels between SZ, BD and HC | | | | | | |
| --- | --- | --- | --- | --- | --- | --- |
| **Inflammatory/immune-related markers^a^** | **SZ  (N = 435)** | **BD  (N = 218)** | **HC  (N = 582)** | ***p*-value^b^** | **Effect size^c^** | **Pairwise comparisons^b^** |
| sTNFR1 | 0.26 (0.16) | 0.25 (0.16) | 0.23 (0.15) | ***p*<0.001** | 0.15 | SZ>HC |
| CXCL16 | 1.22 (0.23) | 1.21 (0.20) | 1.20 (0.21) | ns | - | - |
| OPG | 0.12 (0.12) | 0.15 (0.14) | 0.13 (0.13) | ***p*<0.05** | 0.15 | SZ<BD |
| IL-1RA | 2.36 (0.47) | 2.29 (0.49) | 2.26 (0.48) | ***p*<0.05** | 0.14 | SZ>HC |
| sIL-2R | -0.43 (0.46) | -0.52 (0.33) | -0.55 (0.38) | ***p*<0.001** | 0.21 | SZ,BD>HC |
| Alcam | 1.57 (0.12) | 1.58 (0.11) | 1.58 (0.11) | ns | - | - |
| gp130 | 2.33 (0.10) | 2.32 (0.10) | 2.34 (0.10) | ***p*<0.001** | 0.17 | BD>HC |
| MPO | 2.44 (0.42) | 2.46 (0.44) | 2.46 (0.44) | ns | - | - |
| YKL-40 | 1.58 (0.24) | 1.62 (0.25) | 1.52 (0.21) | ***p*<0.001** | 0.22 | SZ,BD>HC |
| vWF | 1.89 (0.33) | 1.85 (0.36) | 1.86 (0.33) | ns | - | - |
| BDNF | 0.68 (0.30) | 0.69 (0.28) | 0.75 (0.27) | ***p*<0.001** | 0.15 | SZ,BD<HC |
| CatS | 0.71 (0.18) | 0.69 (0.14) | 0.67 (0.15) | ***p*<0.05** | 0.14 | SZ>HC |
| Park7 | 0.70 (0.25) | 0.65 (0.23) | 0.69 (0.26) | ***p*<0.05** | 0.12 | SZ>BD |
| PTX3 | 0.51 (0.40) | 0.47 (0.31) | 0.52 (0.34) | ns | - | - |
| IGFBP4 | 2.19 (0.14) | 2.22 (2.22) | 2.17 (0.13) | ***p*<0.001** | 0.25 | SZ,BD>HC |
| sCD14 | 3.27 (0.10) | 3.28 (0.12) | 3.29 (0.08) | ***p*<0.05** | 0.12 | SZ<HC |
| DKK1 | -0.08 (0.43) | -0.19 (0.34) | -0.06 (0.41) | ***p*<0.001** | 0.19 | BD>HC; SZ<BD |
| Gal3 | 0.44 (0.39) | 0.38 (0.39) | 0.46 (0.41) | ***p*<0.05** | 0.12 | BD<HC |
| MIF | 1.43 (0.54) | 1.38 (0.51) | 1.37 (0.56) | ***p*<0.05** | 0.10 | SZ>HC |
| hs-CRP | 0.36 (0.49) | 0.36 (0.50) | 0.21 (0.47) | ***p*<0.001** | 0.19 | SZ,BD>HC |
| ^a^Mean (standard deviation, SD), log10  ^b^Robust one-way ANOVA and Lincoln post-hoc for pairwise comparison (multiple comparison corrected; WRS2 R-package)^[2]^  ^c^Explanatory measure of effect size, ξ (0.1, 0.3, 0.5, small/medium/large; WRS2 R-package)  Abbreviations: schizophrenia (SZ), bipolar disorder (BD), healthy controls (HC) | | | | | | |

| **Supplementary Table 8.** Cluster comparisons (demographic and clinical data) | | | | | |
| --- | --- | --- | --- | --- | --- |
| **Characteristic** | **Cluster 1** (N = 807)^a^ SZ=161 BD=131 HC=515 | **Cluster 2** (N = 428)^a^ SZ=274 BD=87 HC=67 | **p-value**^b^ | **95% CI**^c^ | **Comparison** |
| Age | 31.43 (9.18) | 33.83 (11.77) | **<0.001** | [-3.69, -1.12] | Cluster 1<Cluster 2 |
| Sex (female) | 426 (53%) | 152 (36%) | **<0.001** | [1.59, 2.58] | Cluster 1>Cluster 2 |
| Education (years) | 13.92 (2.43) | 12.61 (2.51) | **<0.001** | [1.03, 1.61] | Cluster 1>Cluster 2 |
| WASI IQ (2-subtests) | 111.53 (11.42) | 99.84 (13.90) | **<0.001** | [10.15, 13.23] | Cluster 1>Cluster 2 |
| BMI (kg/m²) | 24.56 (3.87) | 27.09 (5.16) | **<0.001** | [-3.13, -1.94] | Cluster 1<Cluster 2 |
| **SMI only** | | | | | |
| PANSS Negative | 10.09 (4.66) | 13.18 (6.22) | **<0.001** | [-3.93, -2.25] | Cluster 1<Cluster 2 |
| PANSS Positive | 7.86 (3.84) | 9.17 (4.41) | **<0.001** | [-1.94, -0.67] | Cluster 1<Cluster 2 |
| PANSS Disorganized | 4.63 (2.06) | 5.47 (2.52) | **<0.001** | [-1.19, -0.49] | Cluster 1<Cluster 2 |
| PANSS Excited | 5.45 (1.93) | 5.55 (2.09) | ns | - | - |
| PANSS Depressed | 8.12 (3.12) | 8.04 (3.23) | ns | - | - |
| YMRS | 4.29 (5.00) | 4.92 (5.20) | ns | - | - |
| GAF Symptom | 50.73 (13.23) | 44.88 (13.20) | **<0.001** | [3.79, 7.89] | Cluster 1>Cluster 2 |
| GAF Function | 50.73 (12.99) | 44.22 (11.43) | **<0.001** | [4.61, 8.42] | Cluster 1>Cluster 2 |
| Age at onset | 23.62 (8.08) | 25.63 (10.06) | ns | - |  |
| Duration of illness (yrs) | 6.14 (6.96) | 7.40 (8.62) | ns | - |  |
| Antipsychotics, DDD | 0.67 (0.85) | 1.09 (1.02) | **<0.001** | [-0.56, -0.27] | Cluster 1<Cluster 2 |
| Antidepressants, DDD | 0.47 (0.78) | 0.50 (0.92) | ns | - |  |
| Antiepileptics, DDD | 0.19 (0.40) | 0.15 (0.38) | ns | - |  |
| Lithium, DDD | 0.09 (0.32) | 0.08 (0.31) | ns | - |  |
| Total, DDD | 1.41 (1.31) | 1.81 (1.43) | **<0.001** | [-0.61, -0.19] | Cluster 1<Cluster 2 |
| ^a^Mean (SD); n (%) ^b^Welch Two Sample t-test; Pearson's Chi-squared test  (Bonferroni corrected, sample characteristics 0.05/5, clinical characteristics 0.05/15) ^c^CI = 95% Confidence Interval of mean difference Abbreviations: schizophrenia (SZ), bipolar disorder (BD), healthy controls (HC) | | | | | |

| **Supplementary Table 9.** Results from inflammatory modulation of iPSC-derived astrocytes and NPCs: SZ donors vs. HC donors | | | | | | |
| --- | --- | --- | --- | --- | --- | --- |
|  | **SZ (N=5)** | **HC (N=5)** | ***t***^b^ | ***p-value***^b^ | **Effect size**^c^ | **BF**^d^ |
| **High IL-1β treatment (acute condition)** | | | | | | |
| **Astrocytes**^a^ | | | | | | |
| YKL-40 | 26.9 (46.0) | 5.1 (2.0) | -1.985 | 0.08 | -1.25 | 1.388 |
| CatS | 0.2 (0.0) | 0.1 (0.5) | 1.126 | 0.29 | 0.71 | 0.716 |
| sTNFR1 | 194.7 (499.5) | 432.6 (94.8) | 0.328 | 0.75 | 0.21 | 0.509 |
| MIF | 1.5 (1.3) | 1.6 (1.1) | 0.039 | 0.97 | 0.02 | 0.492 |
| **NPCs**^a^ | | | | | | |
| YKL-40 | 0.2 (0.3) | 0.47 (0.20) | 1.207 | 0.26 | 0.76 | 0.754 |
| CatS | 0.3 (0.5) | 0.5 (0.4) | 0.563 | 0.59 | 0.36 | 0.543 |
| sTNFR1 | 105.0 (114.0) | 68.0 (31.0) | -1.593 | 0.15 | 1.00 | 0.999 |
| MIF | 0.7 (0.7) | 0.3 (1.1) | 0.339 | 0.74 | 0.21 | 0.510 |
| **Low IL-1β treatment (chronic, low-grade condition)** | | | | | | |
| **Astrocytes**^a^ | | | | | | |
| YKL-40 | 11.7 (19.6) | 3.4 (2.4) | -1.801 | 0.11 | -1.14 | 1.184 |
| CatS | 0.1 (0.0) | 0.2 (0.1) | 1.424 | 0.19 | 0.90 | 0.878 |
| sTNFR1 | 181.4 (160.1) | 266.2 (74.5) | 0.169 | 0.87 | 0.11 | 0.496 |
| MIF | 1.3 (0.1) | 1.0 (0.7) | -0.615 | 0.55 | -0.39 | 0.553 |
| **NPCs**^a^ | | | | | | |
| YKL-40 | 0.1 (0.1) | 0.3 (0.2) | 1.232 | 0.25 | 0.78 | 0.766 |
| CatS | 0.2 (0.1) | 0.2 (0.1) | 0.981 | 0.25 | 0.62 | 0.657 |
| sTNFR1 | 96.0 (37.0) | 64.0 (31.0) | -1.095 | 0.30 | -0.69 | 0.702 |
| MIF | 0.7 (0.6) | 0.7 (1.5) | 0.456 | 0.66 | 0.29 | 0.525 |
| **No IL-1β treatment** | | | | | | |
| **Astrocytes**^a^ | | | | | | |
| YKL-40 | 30.1 (39.3) | 4.6 (1.3) | -2.046 | 0.07 | -1.29 | 1.466 |
| CatS | 0.1 (0.0) | 0.1 (0.0) | 0.792 | 0.45 | 0.50 | 0.596 |
| sTNFR1 | 183.8 (423.8) | 424.0 (70.3) | 0.582 | 0.57 | 0.37 | 0.547 |
| MIF | 1.6 (1.5) | 1.7 (0.4) | -0.299 | 0.77 | 0.19 | 0.506 |
| **NPCs**^a^ | | | | | | |
| YKL-40 | 0.0 (0.0) | 0.1 (0.1) | 2.135 | 0.06 | 1.35 | 1.589 |
| CatS | 0.1 (0.0) | 0.1 (0.0) | 0.375 | 0.71 | 0.24 | 0.514 |
| sTNFR1 | 109.0 (100.0) | 46.0 (56.0) | -1.547 | 0.16 | -0.98 | 0.963 |
| MIF | 0.7 (0.5) | 0.3 (1.1) | 0.226 | 0.82 | 0.14 | 0.500 |
| ^a^Median (IQR)  ^b^Two-sample t-test  ^c^Cohen’s d  ^d^Bayesian t-test, Bayes Factor (BF) computed with default prior (r=0.707)  Abbreviations: schizophrenia (SZ), healthy controls (HC) | | | | | | |

| **Supplementary Table 10.** Results from inflammatory modulation of iPSC-derived astrocytes and NPCs: Low, High and no IL-1β treatment | | | | | | |
| --- | --- | --- | --- | --- | --- | --- |
|  | **No treatment (N=10)** | **Low IL-1β treatment (N=10)** | **High IL-1β treatment (N=10)** | ***p-value***^b^ | **Effect size**^c^ | **BF**^d^ |
| **Astrocytes**^a^ | | | | | | |
| YKL-40 | 5.2 (21.3) | 3.4 (9.3) | 5.2 (19.4) | 0.59 | 0.04 | 0.299 |
| CatS | 0.0 (0.0) | 0.2 (0.1) | 0.2 (0.1) | 0.02 | 0.26 | 3.532 |
| sTNFR1 | 402.2 (251.0) | 254.9 (154.3) | 414.2 (294.8) | 0.11 | 0.15 | 0.922 |
| MIF | 1.6 (1.2) | 1.2 (0.6) | 1.5 (1.2) | 0.02 | 0.24 | 2.746 |
| **NPCs**^a^ | | | | | | |
| YKL-40 | 0.0 (0.1) | 0.2 (0.1) | 0.4 (0.3) | 0.34 | 0.08 | 0.429 |
| CatS | 0.1 (0.0) | 0.2 (0.1) | 0.4 (0.6) | 0.00 | 0.45 | 82.326 |
| sTNFR1 | 70.0 (75.7) | 76.5 (68.7) | 96 (36.5) | 0.52 | 0.05 | 0.324 |
| MIF | 0.7 (0.8) | 0.7 (1.0) | 0.7 (0.9) | 0.63 | 0.03 | 0.287 |
| ^a^Median (IQR)  ^b^one-way ANOVA  ^c^Partial Eta Squared  ^d^Bayesian ANOVA, Bayes Factor (BF) computed with default settings | | | | | | |

| **Supplementary Table 11.** Cognitive characteristics of fibroblast donors | | | | | | | | | | |
| --- | --- | --- | --- | --- | --- | --- | --- | --- | --- | --- |
|  | **SZ**  **(N = 5)** | | | | | **HC (N = 5)** | | | | |
| **Cognitive domains** | **D1** | **D2** | **D3** | **D4** | **D5** | **D1** | **D2** | **D3** | **D4** | **D5** |
| Fine motor speed | - | -0.90 | -4.30 | -0.29 | -0.71 | 0.55 | 0.31 | -0.41 | -0.52 | -0.48 |
| Psychomotor ps | -3.15 | 0.15 | -1.44 | -0.69 | -1.44 | 1.11 | 0.28 | 1.11 | -0.69 | 1.03 |
| Mental ps | - | -0.54 | 0.37 | 0.20 | -0.25 | 1.15 | -0.52 | 0.05 | 0.20 | 0.17 |
| Attention | - | 0.78 | 0.81 | -0.51 | 1.10 | 1.07 | -0.25 | 0.50 | -0.51 | -1.04 |
| Verbal learning | 0.36 | 0.76 | 0.86 | -3.42 | 0.11 | 1.61 | 1.37 | 0.87 | -0.40 | 0.11 |
| Verbal memory | - | 0.69 | 1.00 | -3.07 | -0.17 | 1.07 | 0.41 | 0.69 | 0.41 | 1.00 |
| Semantic fluency | -1.27 | 0.50 | -0.30 | -1.91 | -0.30 | 1.73 | 2.27 | 1.23 | 0.18 | -0.30 |
| Working memory | 0.14 | 1.42 | -0.90 | -2.57 | -0.89 | 1.02 | 0.82 | -1.39 | 0.82 | -0.88 |
| Cognitive control | - | 0.43 | -0.80 | -0.85 | -0.60 | 0.60 | -0.29 | 0.35 | 0.81 | 0.11 |
| Composite | -0.98 | 0.36 | -0.52 | -1.46 | -0.35 | 1.1 | 0.49 | 0.33 | 0.03 | -0.03 |
| ^a^Z-scores calculated based on full N=1235 sample Abbreviations: processing speed (ps), donor (D), schizophrenia (SZ), healthy control (HC) | | | | | | | | | | |

1. Patrick Mair and Rand Wilcox, ‘Robust Statistical Methods in R Using the WRS2 Package’, *Behavior Research Methods*, 52.2 (2020), 464–88 (p. 2) <https://doi.org/10.3758/s13428-019-01246-w>.
2. Patrick Mair and Rand Wilcox, ‘Robust Statistical Methods in R Using the WRS2 Package’, *Behavior Research Methods*, 52.2 (2020), 464–88 (p. 2) <https://doi.org/10.3758/s13428-019-01246-w>.
