## Supplementary material for "Cognitive and inflammatory heterogeneity in severe mental illness: Translating findings from blood to brain": Supp_Res

**Supplementary Results**

1. **Canonical correlation analysis (CCA)**

The CCA revealed three significant patterns of covariation between cognitive domains and peripheral marker levels using the Wilk’s Lambda test statistic (Mode_1_: r=0.41, *p*<0.001, Mode_2_: r=.22, *p*<0.001, Mode_3_: r=0.20, *p*=0.01), while all modes were significant via permutation (See Fig. S3). Cross-validation showed that only the first mode performed relatively well on unseen data (mean_training_=0.41, mean_test_=0.36), whereas all other modes had substantially lower canonical correlation in the test sets compared to the full sample (i.e. mode 2: mean_training_=0.23, mean_test_=0.06; mode 3: mean_training_=0.21, mean_test_=0.07). Due to poor performance of other modes in the out-of-sample cross-validation, suggestive of low generalizability, we only considered the first mode. The jack-knife procedure showed that small perturbations did not cause large variations in the canonical loadings, suggesting robust loadings even in the presence of outliers (Fig. S4).

2. **Hierarchical Clustering analysis**

Ward’s linkage method showed the highest agglomerative coefficient (0.99). The silhouette index was maximized (0.39) for a 2-cluster solution (Fig. S5). Data simulation resulted in a significant silhouette index (*p*=0.005), indicating a rejection of the null hypothesis that the data comes from a single Gaussian distribution (Fig. S6). The stability analysis suggested robust cluster assignment for cluster 1, with 76% overlap following bootstrapping, but was slightly less robust for cluster 2 (65% overlap).
