## Supplementary material for "Cognitive and inflammatory heterogeneity in severe mental illness: Translating findings from blood to brain": Supp_Figs

**Supplementary Figures**

1. **Cognitive domains from test battery I and II**

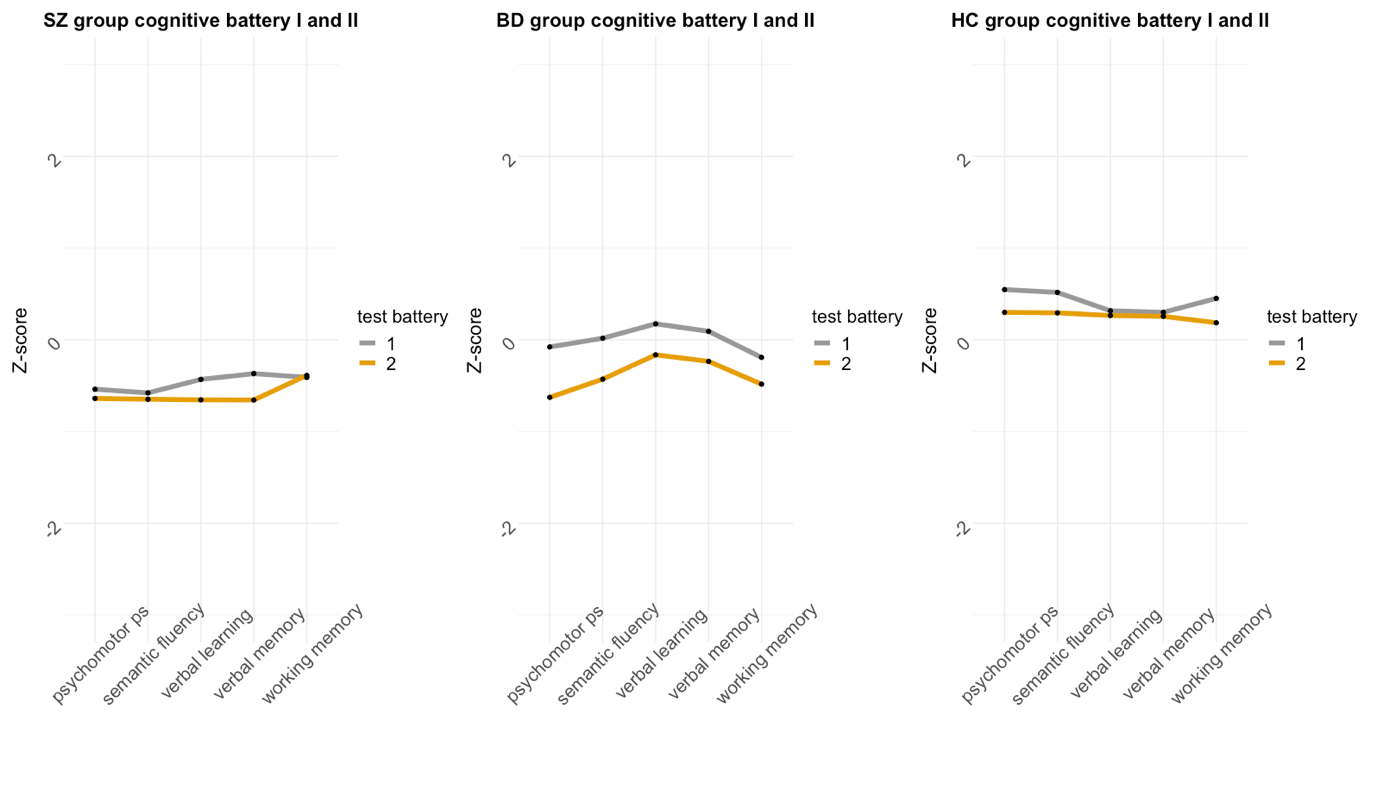

**Supplementary Fig. 1** Overview of Z-scores from the cognitive domains using different tests from different cognitive test batteries, stratified by group. Cognitive tests from battery II seem to be slightly more sensitive than the tests in battery I. However, MANOVA analyses revealed no significant difference in each group between the scores in battery I and battery II when correcting for multiple comparisons (*p*>0.01 for all comparisons). Supplementary table X also show similar variance across these cognitive tests.

Abbreviations: schizophrenia (SZ), bipolar disorder (BD), healthy controls (HC)

2. **MICE outputs**

**A**

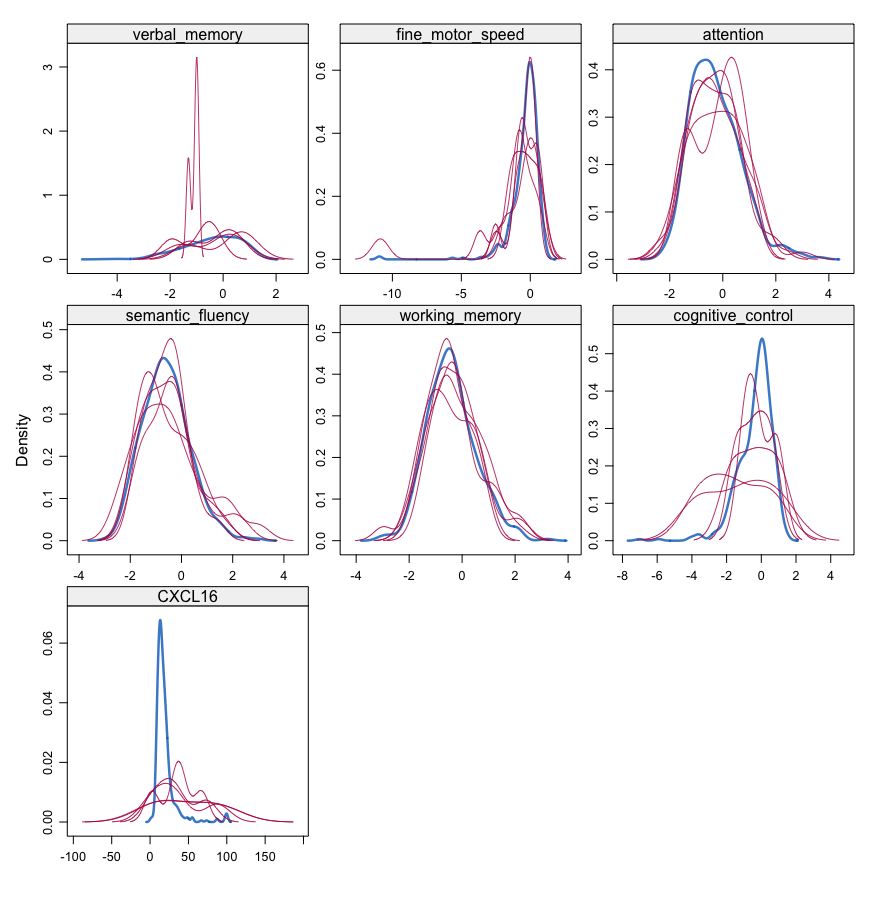

**B**

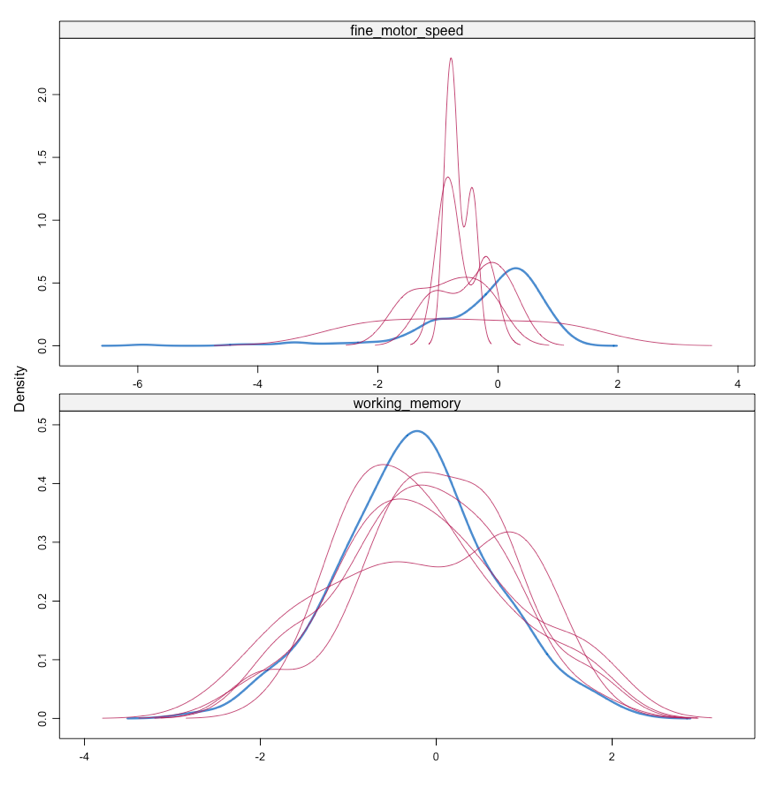

**C**

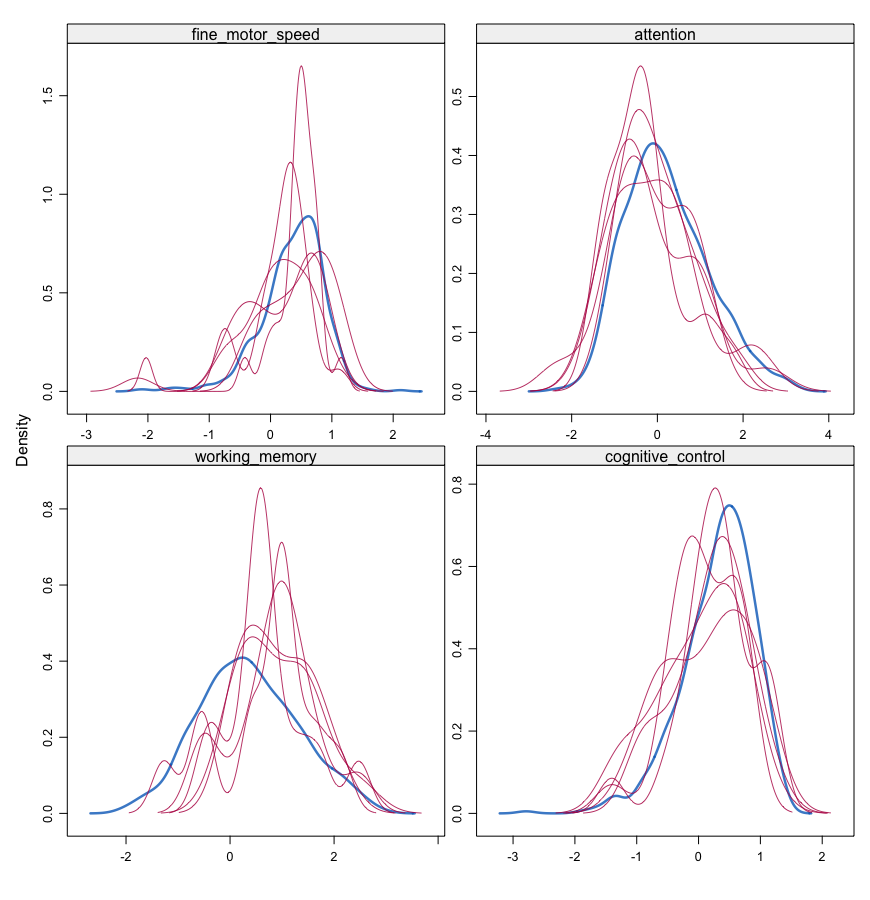

**Supplementary Fig. 2 (A-C)** Shows the density of the imputed data for each imputed dataset (in magenta) and the observed data (blue). Imputations (*m*=5) with similar distributions as observed data is the best fit, we selected imputation dataset 1. MICE imputation was completed separately for A) the SZ group, B) the BD group, and C) the HC group.

3. **Permutation analysis (CCA)**

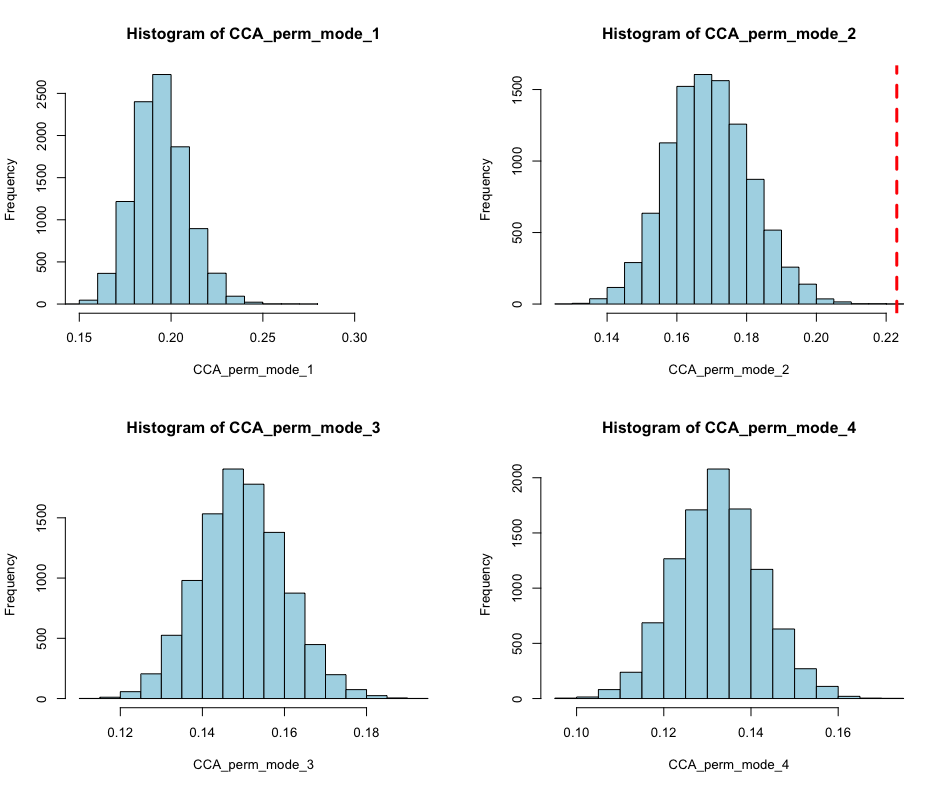

**Supplementary Fig. 3** Shows the null distribution from the permutation analysis (four first modes), where 8/9 modes were significant with permutation. However, only the first mode showed performed well on unseen data from 10-fold cross-validation. Only the first mode was considered in follow-up analyses.

4. **Stability of canonical loading scores (CCA)**

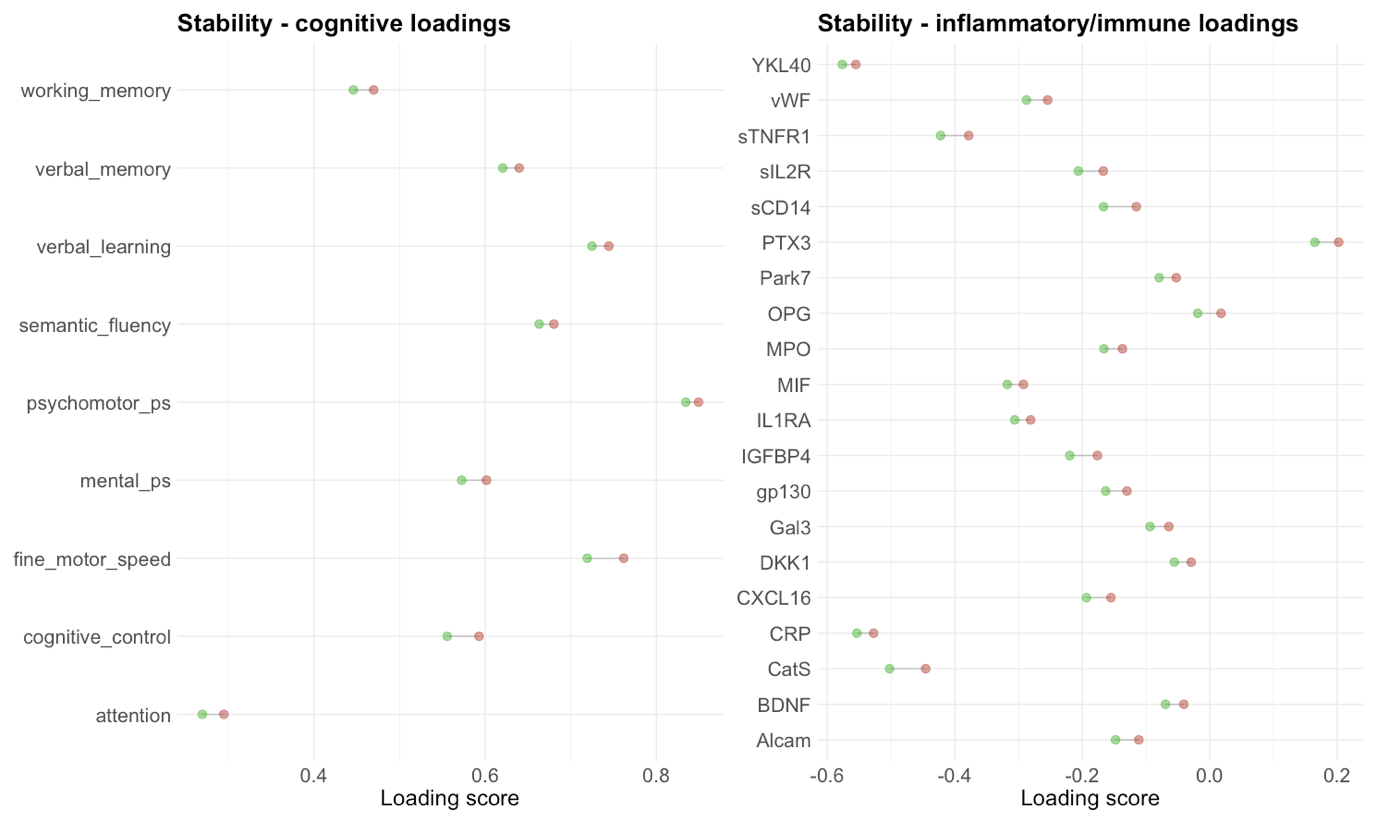

**Supplementary Fig. 4** Results from the stability analysis based on Dinga et al., (2019) with available R code at github (<https://github.com/dinga92/niclin2019-biotypes>). Both the cognitive and inflammatory/immune marker loadings showed robust canonical variate loadings when running the CCA and leaving one participant out of the analysis.

5. **Hierarchical clustering: Evaluation of n clusters**

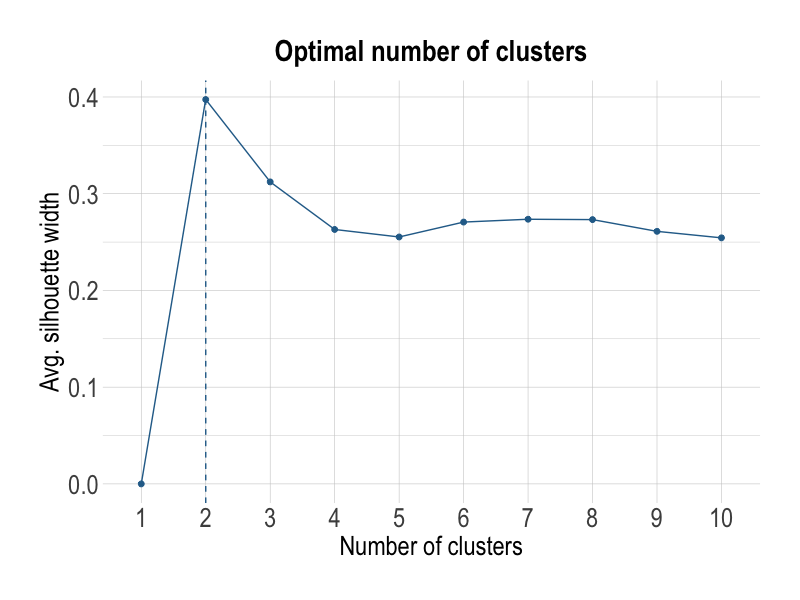

**Supplementary Fig. 5** Evaluation of n clusters for hierarchical clustering of canonical variates using the average silhouette index. The average silhouette index was maximized for a 2-cluster solution.

6. **Hierarchical clustering: Significance test**

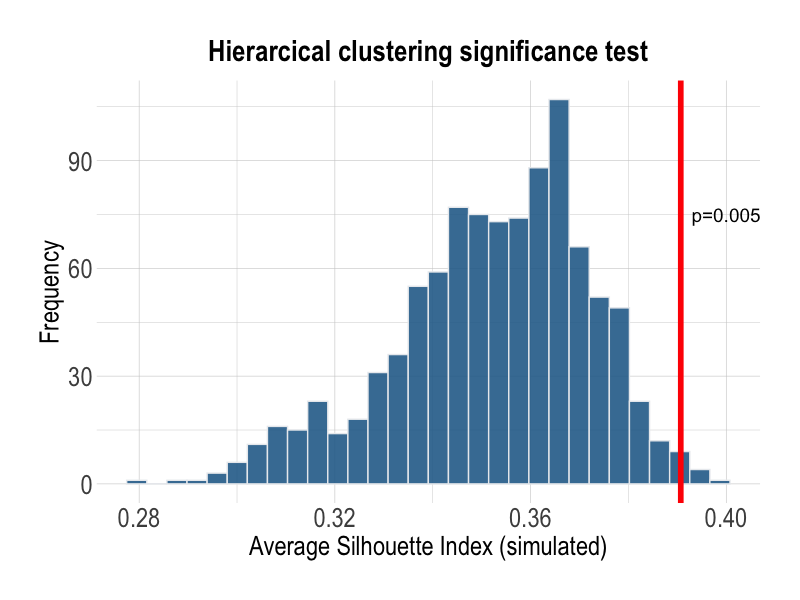

**Supplementary Fig. 6** Shows null distribution and results from significance test of the average silhouette index. A significant result (*p*<0.005) indicates we can reject the null hypothesis (i.e. data comes from a single normal Gaussian distribution).

7. **Correlation among candidate marker genes and inflammatory genes in postmortem samples (CMC)**

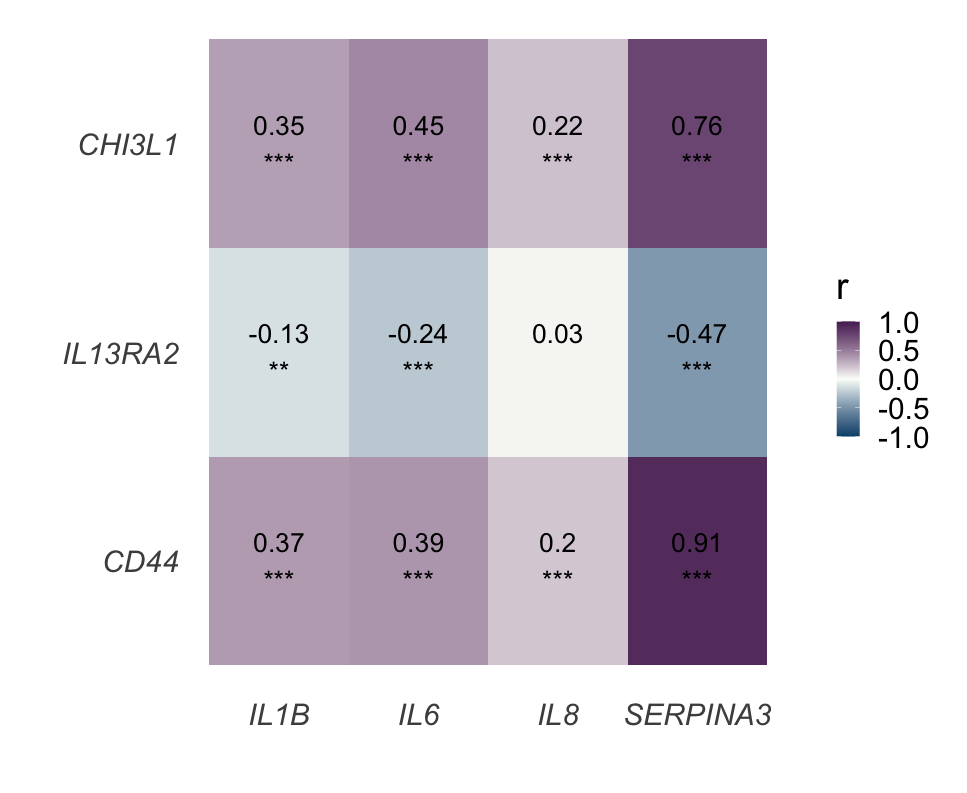

**Supplementary Fig. 7** Shows Pearson correlation between candidate marker genes and inflammatory genes used to generate high-low inflammation subgroups in dlPFC postmortem samples from the CMC.
