## Supplementary material for "Cognitive and inflammatory heterogeneity in severe mental illness: Translating findings from blood to brain": Supp_Methods

**Supplementary Methods**

1. **Generation of iPSC-astrocytes and NPCs**

The generation and characterization of iPSCs and differentiation of iPSC-derived astrocytes were done as described previously [1]. Briefly, fibroblasts isolated from control (n=5) and patient donors (n=5) were reprogrammed using Sendai virus, transduced with the CytoTune™-iPS 2.0 Sendai Reprogramming Kit (Thermo Fisher, Waltham, MA, USA) containing KOS (Klf4, Oct4, Sox2), Nanog, and c-Myc reprogramming factors. Each iPSC line was subjected to rigorous quality control at The Norwegian Core Facility for Human Pluripotent Stem Cells at the Norwegian Center for Stem Cell Research including phenotyping, regular monitoring of morphology, and pluripotency marker expressions. Karyotyping to assess the chromosomal integrity of all the derived iPSC lines was performed at passage 15 for verification of authenticity and normality (KaryoStat Karyotyping Service, Thermo Fisher). Karyotyping test results and iPSC phenotyping data have been published previously [2], and are available upon request.

For astroglia and NPC differentiation, iPSC colonies at passages 24-25 were transferred to 6-well tissue culture plates in Neural Maintenance Medium (NMM) consisting of 50% DMEM/F12 and 50% Neurobasal Medium (both from Thermo Fisher) supplemented with 0.5% (v/v) N2, 1% (v/v) B27 (both from Invitrogen, Carlsbad, CA, USA), 5 μg/ml human insulin, 40 ng/ml triiodothyronine (T3), 10 μM β-mercaptoethanol, 1.5 mM L-glutamine, 100 μM NEAA, 100 U/ml penicillin, and 100 μg/ml streptomycin (all from Sigma-Aldrich, St. Louis, MO, USA). 20 ng/ml EGF and 4 ng/ml bFGF (both from Peprotech, Rocky Hill, NJ, USA) were added to the cultures. After 1 day, non-adherent embryoid bodies (EBs) formed in the cultures and were fed daily with NMM containing T3, EGF, and bFGF (as above) until day 3. On day 3, 10 μM all-trans retinoic acid (ATRA, Sigma) was added to the medium and EBs were washed (1x, gently in NMM medium) and plated on Geltrex-coated plates (Thermo Fisher). After this step, cultures were fed with ATRA + T3 + growth hormone supplemented NMM medium daily until day 10. On day 10, neurorosette formation was monitored by light microscopy, and NES (Nestin) and PAX6 positivity was checked by qPCR. Cultures were passaged using the cell dissociation agent Accutase (Sigma) following the recommended protocol. From this point on, regular passaging was done at confluence and cultures were seeded on Geltrex-coated 6-well plates. On day 10, the medium was changed to NMM + 40 ng/ml T3 + 20 ng/ml EGF. On day 18, after the formation of neural progenitor cells (NPCs), the medium was changed to NMM supplemented with B27 without vitamin A (Invitrogen) + 20 ng/ml EGF + 40 ng/ml T3, and cultures were fed using the same medium composition until day 40. Samples were continuously collected during the differentiation process at days 0 (iPSC), 7, 14, 21, 30, and 40. The mRNA expression of the neural progenitor markers PAX6 and NES, and the astrocyte-specific markers GFAP (Glial fibrillary acidic protein), S100B (S100 calcium-binding protein B), AQP4 (Aquaporin 4), SLC1A2 and SLC1A3 (Solute Carrier Family 1 Member 2 and 3), FABP7 (Fatty acid-binding protein 7), ALDH1L1 (Aldehyde Dehydrogenase 1 Family Member L1), and ALDOC (Aldolase C) was monitored by qPCR using a custom-designed TLDA gene array card (Thermo Fisher). Cells were stained with anti-GFAP, anti-S100B, and anti-AQP4 antibodies (all from Abcam, Cambridge, UK) and were analyzed using fluorescence microscopy on day 40. Fully differentiated iPSC-derived astrocytes expressing the relevant astroglia-specific markers were used for functional studies on day 40.

IPSCs from CTRLs and patients were differentiated into neural precursor cells (NPCs) and phenotyped/characterized as described previously [3]. First, iPSCs on Geltrex-coated plates at a 15–20% density were differentiated to neuroepithelium with rosette formation in A-DMEM/F12, 1% penicillin streptomycin, 1% Glutamax and 1% N2 with the addition of SB431542, LDN193189 and XAV-939 (dual SMAD inhibition and Wnt inhibition). At day 7, rosette-like structures were isolated with Rosette Selection Reagent (Stemcell technologies) and 9x10^5^cells/well were plated on laminin coated plates. NPCs were then cultured in A-DMEM/F12, 1% P/S, 1% Glutamax, 1% N2 and 0,4% B27 supplemented with FGF2 at 2,5ng/ml changing the medium every other day during 5-6 days. NPCs were then passaged and cultured in A-DMEM/F12, 1% P/S, 1% Glutamax, 1% N2, 0,1% B27 in hypoxic conditions (3% O2) on laminin coated 6-well plates. Basal medium was equilibrated in the incubator O/N and FGF2 was added fresh daily to the cells to a final concentration of 10ng/ml. Cells were passaged with accutase (Sigma) every 4-5 days.

2. **R-packages used for visualization and statistical analyses**

- For visualization the R-package “ggplot2” was applied [4].
- Multiple Imputation by Chained Equations (MICE) was applied using the R-package “mice” [5].
- For permutation-based t-tests (sample & clinical characteristics) the R-package “rcompanion” [6] was used.
- The CCA and permutation test was implemented using R-packages “candisc” [7] and “rsample” [8].
- The 10-fold cross-validation procedure was implemented using the R-package “caret” [9].
- Clustering was performed using R-packages “cluster” [10], “dendextend” [11] and “factoextra” [12].
- Clustering stability was assessed using the R-package “fpc” (flexible procedures for clustering) [13].
- Bayesian t-tests were assessed using the R-package “BayesFactor” [14].
- Correlation analyses were run using the R-package “Hmisc” [15]

3. **CCA: Permutation, cross-validation, and stability**

The significance of CCA modes was assessed using permutation testing, and the traditional Wilks Lambda test statistic. In each permutation, CCA was repeated by randomly shuffling the rows of biomarker data matrix such that they no longer corresponded to rows in the cognitive data matrix, thereby breaking the association between the two datasets. We performed 10000 permutations, generating a null distribution of canonical correlations, controlling for multiple comparisons across possible significant modes. When assessing this statistic, we identified three significant modes, although permutation testing identified all modes as significant. However, only the first mode was considered further due to low generalizability by cross-validation of all other modes.

It is recommended to perform additional dimension reduction if the subject-variable-ratio (SVR) in CCA is very low, i.e. n variables is very large (>50) [16]. Due to the high SVR in our study, we did not perform dimension reduction prior to CCA. The recommended sample size for detecting reliable effects in CCA was met, as the recommendation is 20 times as many cases as variables (N_variables_=29, minimal sample size required 580; Our N_sample_=1235) [17, 18].

The stability analysis (leave-one-out procedure) involved generating a distribution of the canonical loadings for each resample which was plotted to evaluate loading stability. A large variation in the canonical loadings under small changes indicates the CCA model is unstable [19]. The stability analysis can help identify whether outliers have significant impact on the canonical loadings. In our case the stability analysis indicated stable loadings under the leave-one-out procedure, as can be observed in Fig. SX.

4. **Hierarchical clustering: Significance and stability analyses**

It has been noted that an observed maximum silhouette index for n-clustering solution does not provide evidence for the presence of n-clusters in a given dataset. Therefore we followed the recommended data simulation procedure by Dinga and colleagues [19], which tests the null hypothesis that the data comes from a single normal Gaussian distribution (i.e. there are no clusters in the observed data). The procedure involves estimating a covariance matrix based on the cognitive and peripheral biomarker canonical variates (loading scores) used in the hierarchical clustering analysis. In each simulation (n=1000), random samples were drawn from a normal distribution based on the covariance matrix. Hierarchical clustering was then performed on each random sample, and the maximum silhouette index was calculated. This generated an empirical null distribution of the silhouette index, from which the *p*-value was defined as the number of times the silhouette index was smaller in the null distribution compared to the observed data. A significant result indicates we can reject the null hypothesis that the data comes from a single Gaussian normal distribution.

The clustering assignment was evaluated under small perturbations of the data using bootstrapping. The cognitive and peripheral biomarker canonical variate loading scores were resampled using bootstrap, and the Jaccard similarity index was used to compare the similarities of the original clustering solution to the most similar clusters in the resampled data. The mean Jaccard similarity index over these similarities was used as an index of the stability of the clustering solution. A Jaccard similarity index <=0.5 is considered unstable, 0.6-0.7 indicate presence of a pattern in the data, and >=0.75 indicate robust clusters [20].
